# Neonatal outcomes and indirect consequences following maternal SARS-CoV-2 infection in pregnancy: A systematic review

**DOI:** 10.1101/2022.05.20.22275313

**Authors:** Sarah Sturrock, Shohaib Ali, Chris Gale, Cheryl Battersby, Kirsty Le Doare

**Author notes:** **Contact details for corresponding author:** Sarah Sturrock, Institute for Infection & Immunity, Paediatric Infectious Diseases Research Group, St George’s, University of London, Cranmer Terrace, London, SW17 0RE, 020 8672 9944.

## Abstract

**Objectives:** Identify the association between maternal SARS-CoV-2 infection in pregnancy and individual neonatal morbidities and outcomes, particularly longer-term outcomes such as neurodevelopment.

**Setting:** Case-control and cohort studies from any location published after 1st January 2020, including pre-print articles.

**Participants:** Neonates born to pregnant women diagnosed with a SARS-CoV-2 infection at any stage during pregnancy, including asymptomatic women.

**Primary and secondary outcome measures:** Neonatal mortality and morbidity, including preterm birth, Caesarean delivery, small for gestational age, admission to neonatal intensive care unit, level of respiratory support required, diagnosis of culture-positive sepsis, evidence of brain injury, necrotising enterocolitis, visual or hearing impairment, neurodevelopmental outcomes, and feeding method. These outcomes were selected according to a Core Outcome Set developed between health professionals, researchers and parents.

**Results:** The search returned 3234 papers, from which 204 were included with a total of 45,646 infants born to mothers with SARS-CoV-2 infection during pregnancy across 36 countries. We found limited evidence of an increased risk of some neonatal morbidities, including respiratory disease. There was minimal evidence from low-income settings (1 study) and for neonatal outcomes following first trimester infection (17 studies). Neonatal mortality was very rare. Preterm birth, neonatal unit admission and small for gestational age status were more common in infants born following maternal SARS-CoV-2 infection in pregnancy in most larger studies.

**Conclusions:** There is limited data on neonatal morbidity and mortality following maternal SARS-CoV-2 infection in pregnancy, particularly from low-income countries and following early pregnancy infections. Large, representative studies addressing these outcomes are needed to better understand the consequences for babies born to women with SARS-CoV-2 in pregnancy.

**Trial registration:** PROSPERO ID: CRD42021249818

**Strengths and limitations:**

- Inclusion of studies of both asymptomatic and symptomatic SARS-CoV-2 infections at any point in pregnancy to maximise generalisability of findings
- Focus on neonatal outcomes, as opposed to purely obstetric outcomes, to accurately quantify neonatal morbidity
- Study is limited by available data; important data gap in low-income settings

## Introduction

Pregnant women have been treated as an ‘at risk’ group for severe disease during the SARS-CoV-2 pandemic[1]. Initial evidence suggested that infection with SARS-CoV-2 in pregnancy was associated with severe obstetric morbidity[2], including higher rates of preterm birth, pre-eclampsia, and Caesarean delivery [3][4]. Early case reports suggested that vertical transmission was possible, although rare[2][5][6][7][8][9]. However, increasingly, research indicates that neonatal infections are mostly mild[10], suggesting that the risk to neonates from maternal infection is more likely to be as a result of the indirect effects of being born to a mother with SARS-CoV-2 infection, rather than from perinatal or postnatal infection with SARS-CoV-2. Other viral infections, such as Zika virus, in early pregnancy have been associated with adverse neurodevelopmental outcomes[11][11]; however, the neurodevelopmental impact of maternal SARS-CoV-2 in pregnancy is unclear.

Previous reviews of neonatal outcomes from maternal SARS-CoV-2 infections have been limited by the quality of evidence available, with many early studies consisting of case reports and case series. As larger, population-based or national studies emerge, an opportunity has arisen to examine neonatal outcomes following maternal infection in greater detail, including longer-term outcomes. In this systematic review, we summarise current evidence on neonatal outcomes after maternal SARS-CoV-2 infection in pregnancy, aiming to quantify the association with specific neonatal morbidities and longer-term outcomes that will be important to families.

## Methods

The review protocol was pre-registered and is available with PROSPERO (17th May 2021, ID CRD42021249818).

### Eligibility criteria

We included peer-reviewed publications of case-control and cohort studies. Pre-print articles identified from relevant living systematic reviews were included. We excluded studies of overlapping populations, identified by hospital, date of study period, and number of participants. Pre-print articles were identified as reporting duplicate populations by the same means. We included studies of the babies of pregnant women with a diagnosis of SARS-CoV-2 during pregnancy. A diagnosis of SARS-CoV-2 was defined as positive polymerase chain reaction (PCR) testing, lateral flow/rapid antigen testing, or locally accepted clinical criteria in order to enable inclusion of studies early in the pandemic or in resource-limited settings where PCR testing may not have been widely available. We included studies of SARS-CoV-2 diagnoses at any stage during pregnancy. We included studies diagnosing SARS-CoV-2 infection using serology only for studies including participants in the first 9 months of 2020, with the assumption that these participants would mostly have contracted their primary SARS-CoV-2 infection during pregnancy. In case-control studies, we included any study with a comparison group of pregnant women without any diagnosis of SARS-CoV-2 during pregnancy. We included studies published after 1 ^st^ January 2020, although studies published after this date but including data from prior to 1^st^ January 2020 were also included. No language or geographic restrictions were applied.

We included studies describing any of the following infant outcomes: preterm birth (<37 weeks gestation), small for gestational age (<10^th^ centile birthweight for gestational age on appropriate neonatal growth charts), low birthweight (<2500g), admission and length of stay in neonatal unit, level and duration of respiratory support, diagnosis of culture-positive sepsis during neonatal admission, evidence of brain injury (including seizures, abnormal brain imaging, or diagnosis of hypoxic ischaemic encephalopathy)[12], necrotising enterocolitis, other gastrointestinal disease, visual or hearing impairment, quality of life, neurodevelopmental outcomes, exclusive breastfeeding, and all cause infant mortality. Selection of neonatal outcomes were informed by a Core Outcome Set developed with health professionals, parents, and researchers[13].

### Search process

MEDLINE, Embase, Global Health, WHOLIS and LILACS databases were searched (see Appendix 1 for search terms used). The LILACS database was searched for all papers relating to “SARS-CoV-2”, “covid” and “coronavirus”, owing to its differing search functionality from the other databases. The last search was completed on 28^th^ July 2021.

Results were uploaded to the Rayyan QCRI platform (Rayyan – a web and mobile app for systematic reviews, 2016[14]), and duplicates removed using the duplicate removal tool available on this platform. All titles were screened independently by 2 reviewers (SS and AS), and subsequently abstracts screened by both. Where there was disagreement, the title/abstract was screened by a third reviewer (CG).

Data were extracted into Microsoft Excel (Version 2201) by SS or SA using a proforma with the outcomes described above, study type and dates, location, participant definition and numbers, and method of SARS-CoV-2 diagnosis. Any outcome data not reported was assumed not to have been collected as part of the study. Pregnancies were assumed to be singleton pregnancies unless otherwise specified. A modified Newcastle-Ottawa Scale[15] was used for assessment of study quality, with studies scoring 4 and above (out of a possible 11) deemed as eligible for inclusion. Statistical analysis was completed using Microsoft Excel, SPSS (IBM SPSS Statistics for Macintosh, Version 25, 2017[16]) and R (R Studio Version 2021.09.01[17]), including calculation of proportion of infants in each study with each outcome, and descriptive statistics of rates of outcomes identified. Weighted means were calculated by dividing the number of infants included in each study by the total number of infants included in the review to find a weighting factor. Each outcome rate was then multiplied by that study’s weighting factor, and all the results summed to find the overall weighted mean. Independent sample Kruskal-Wallis tests were used to determine whether there was a significant difference in outcome rates between country income levels as defined by the World Bank[18]. Forest plots were created using R[17], using a random effects model only. Further meta-analysis was not performed due to heterogeneity in study populations and outcome reporting. Results are reported according to PRISMA guidelines.

### Patient and public involvement

Patients and the public were not directly involved in the design of this study. However, this study seeks to address some of the knowledge gaps raised by expectant families as part of an online survey of women pregnant or breastfeeding during the COVID-19 pandemic[19].

### Ethics Approval

Ethical approval was not required for this study, as it involved only retrieval and synthesis of data from previously published studies.

## Results

### Search results

3234 papers were identified from the literature search after duplicates were removed. A total of 204 papers were deemed as eligible for inclusion. Of these, 37 papers were case-control studies, and 167 were cohort studies (see Figure 1 for PRISMA summary of study selection process). A total of 36 countries were represented, with an additional 6 international papers. 118 studies were from high-income countries, and only 1 from a low-income country[20]. Study periods ranged from the 8^th^ December 2019 to 23^rd^ February. Across all studies, a total of 838,743 pregnancies and 786,884 live births were studied, of which 57,059 mothers had received a diagnosis of SARS-CoV-2 infection in pregnancy and had given birth to 45,646 babies. The majority of women had SARS-CoV-2 in their third trimester of pregnancy, with 17 (8.3%) studies including participants in the second trimester (2-49% of total participants in each study), and 20 (9.8%) including first-trimester participants (1-51% of total participants in each study). 76% of studies used PCR testing alone to identify cases of SARS-CoV-2. Details of included studies can be found in Table 1, and a full results table is available in Appendix 2. The range of bias assessment scores according to the Newcastle-Ottawa scale were 4-8, with a median score of 6.

**Table 1:** Study demographics

|  |  | Number of studies | Number of participants |
| --- | --- | --- | --- |
| Study type | Case-control | 37 | 793680 |
|  | Cohort | 167 | 45063 |
| Income group | High | 118 | 809562 |
|  | Upper middle | 56 | 15027 |
|  | Lower middle | 24 | 7174 |
|  | Low | 1 | 137 |
| Trimester of infection | First trimester included | 20 | 2212 |
|  | Second trimester included | 17 | 2141 |

**Figure 1:**
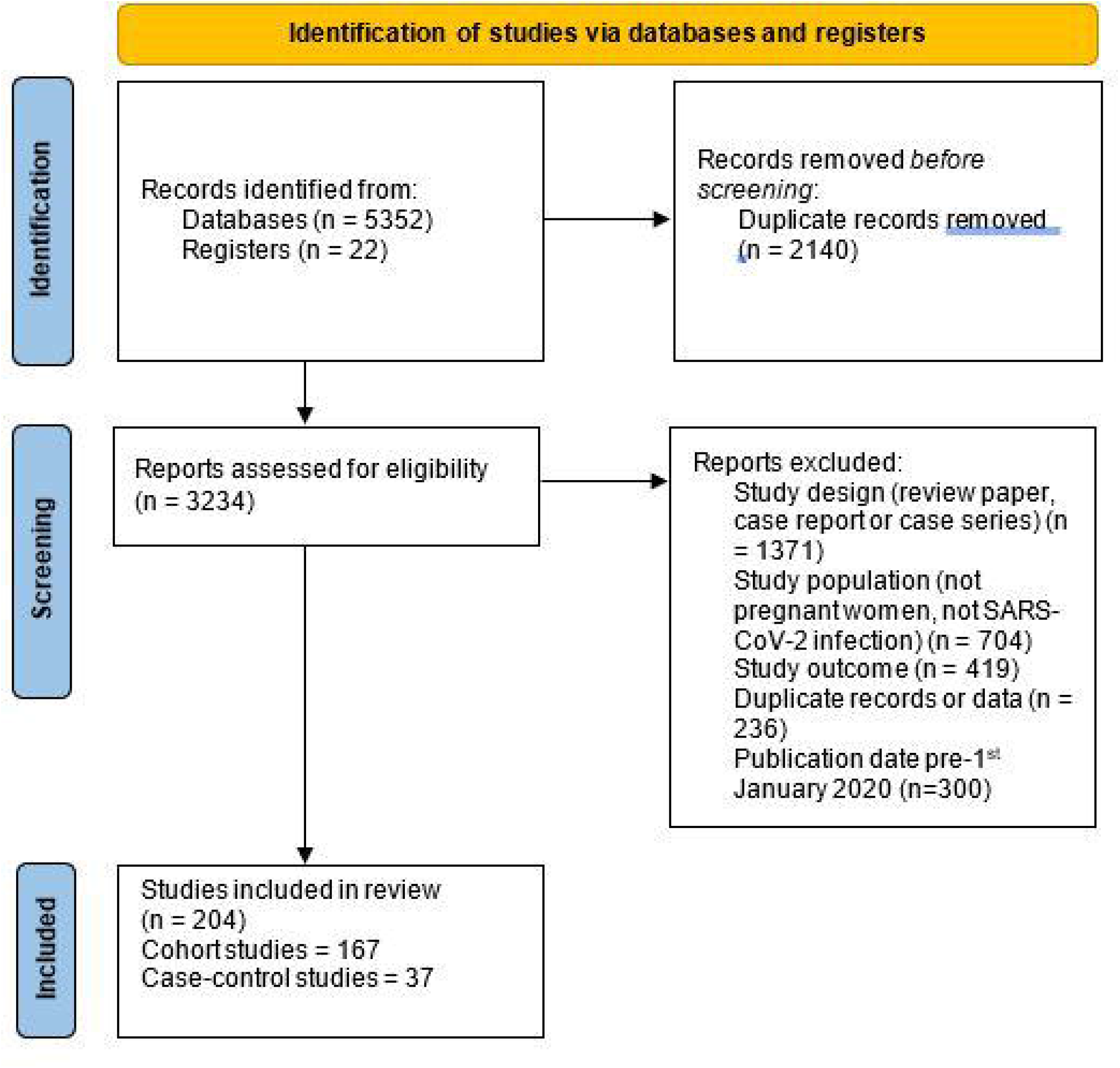
PRISMA study selection flow chart

### Neonatal morbidity

Of the included studies, neonatal outcomes were less commonly reported than obstetric outcomes. Need for admission to a neonatal unit was the most frequently reported outcome, with data extracted for 761,489 infants respectively (97.2% of included infants overall). However, neonatal outcomes such as need for non-invasive respiratory support, neurological disease, sepsis and necrotising enterocolitis were only reported in a minority of infants (<95,000) and studies included in this review.

The weighted mean rate of admission to a neonatal unit for babies born to mothers infected with SARS-CoV-2 was 11%, although it was not clear in some studies how many of these admissions were for isolation purposes as opposed to clinical need. 8 of the 19 case-control studies reporting neonatal unit admission rates found a significant association between neonatal unit admission and maternal infection (including 432,512 infants, in comparison to 306,407 infants included in studies finding no association, see table 2 and figure 2). The need for non-invasive respiratory support amongst babies born to mothers with SARS-CoV-2 was reported in 94,970 infants (weighted mean rate 1%, see table 3). Neurological disease (reported in 89,337 infants, range 0-7%, weighted mean rate 0.2%), NEC (reported in 88,773 infants, weighted mean rate 0.02%), and confirmed bacterial infection (reported in 93,547 infants, range 0-7%, weighted mean rate 0.09%) were all reported in a minority of studies. Few case-control studies reported on neonatal morbidity in detail, with only 2 studies of 88,238 infants examining the need for respiratory support, gastrointestinal disease, neurological disease, and sepsis. One small case-control study of 79 infants found maternal SARS-CoV-2 infection to be associated with neurological morbidity (specifically, seizures), affecting 1 (7%) of the exposed infants and none of those non-exposed[21]. One large study of 88,159 infants finding an increased risk of need for respiratory support in babies born to infected mothers found that this association may be explained by prematurity[22]. No study controlled for prematurity in assessing the association between maternal infection and neurological morbidity.

**Table 2:** results of case-control studies

|  |  | Studies finding significant association |  | Studies not finding significant association |  |
| --- | --- | --- | --- | --- | --- |
|  |  | Number of studies | Number of participants in studies | Number of studies | Number of participants in studies |
| Birth outcomes | Premature delivery (<37 weeks) | 10 | 648804 | 16 | 9807 |
|  | Small for gestational age | 1 | 219 | 10 | 648318 |
|  | Low birthweight | 1 | 2130 | 1 | 110 |
| Neonatal outcomes | Admission to neonatal care | 8 | 432512 | 11 | 306407 |
|  | Need for non-invasive respiratory support | 2 | 88238 | 0 | 0 |
|  | Need for mechanical ventilation | 2 | 88238 | 0 | 0 |
|  | Neurological disease | 1 | 79 | 1 | 88159 |
|  | Necrotising enterocolitis | 0 | 0 | 0 | 0 |
|  | Other gastrointestinal disease | 0 | 0 | 0 | 0 |
|  | Sepsis | 0 | 0 | 2 | 88214 |
| Infant outcomes | Hearing impairment | 2 | 191 | 0 | 0 |
|  | Developmental outcomes | 0 | 0 | 0 | 0 |
|  | Breastfeeding | 2 | 145 | 2 | 88422 |
|  | Infant or neonatal death | 0 | 0 | 10 | 96688 |

**Table 3:** Results from all COVID-19 positive pregnancies (cohort studies and case-control studies)

|  |  | Number of studies reporting | Number of infants included | Weighted mean | Range |
| --- | --- | --- | --- | --- | --- |
| Birth outcomes | Premature delivery (<37 weeks) | 165 | 782612 | 13.8% | 0-81% |
|  | Small for gestational age | 55 | 753945 | 4.0% | 0-44% |
|  | Low birthweight | 25 | 5108 | 1.0% | 0-50% |
| Neonatal outcomes | Admission to neonatal care | 118 | 761489 | 11.0% | 0-100% |
|  | Need for non-invasive respiratory support | 27 | 94970.00 | 1.0% | 0-80% |
|  | Need for mechanical ventilation | 27 | 94454 | 0.4% | 0-20% |
|  | Neurological disease | 13 | 89337 | 0.2% | 0-7% |
|  | Necrotising enterocolitis | 10 | 88773 | 0.0% | 0-22% |
|  | Other gastrointestinal disease | 6 | 360 | 0.0% | 0-5% |
|  | Sepsis | 15 | 93547 | 0.1% | 0-7% |
| Infant outcomes | Hearing impairment | 4 | 233 | 0.1% | 0-31% |
|  | Developmental outcomes | 2 | 339 | 0.0% | 0-64% |
|  | Breastfeeding | 38 | 96174 | 12.0% | 0-100% |
|  | Infant or neonatal death | 99 | 117613 | 0.4% | 0-18% |

**Figure 2:**
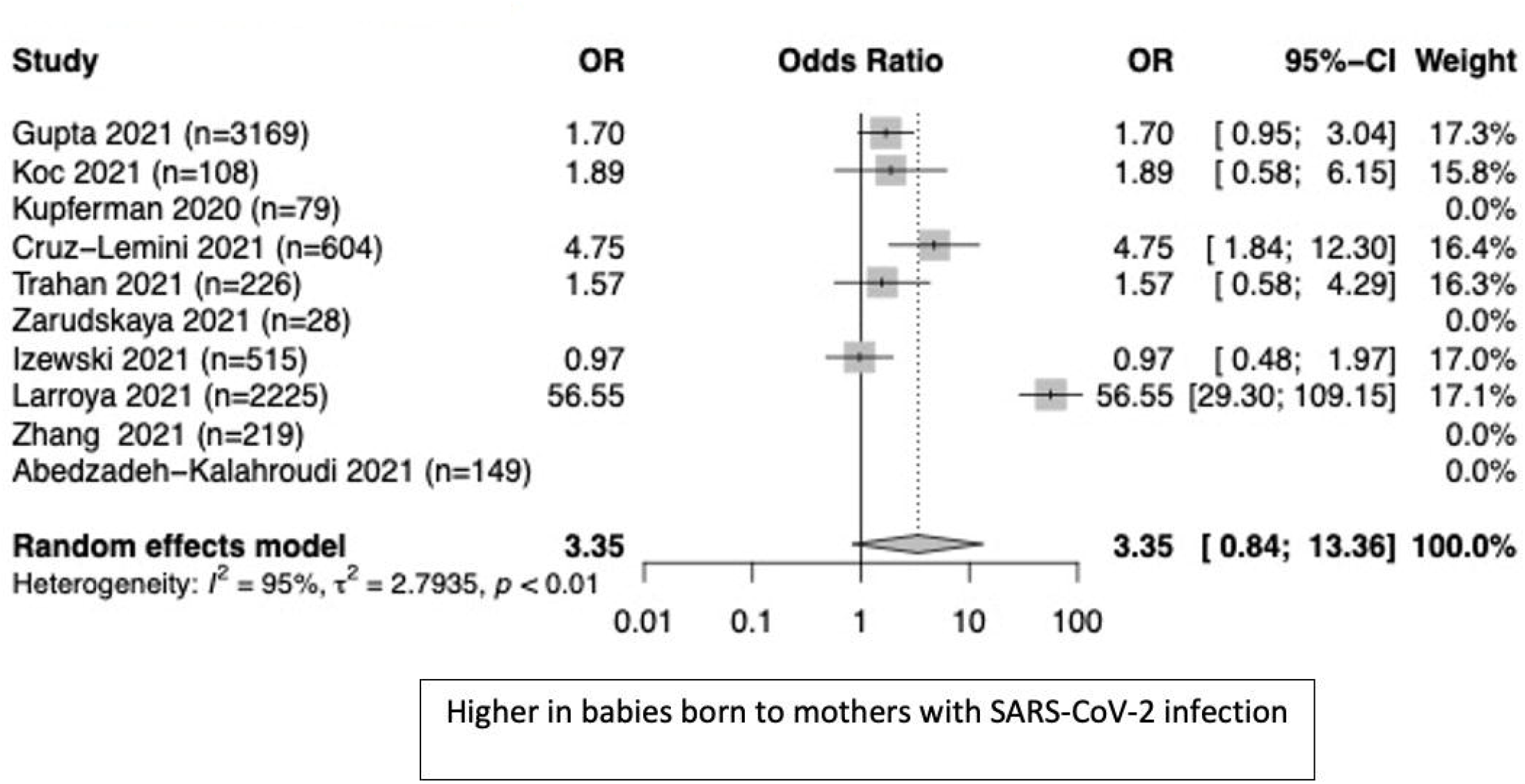
Forest plot for NICU admission

### Birth outcomes

Method of delivery was reported in 184 studies (including 784,395 births), with a weighted mean of 38% of births occurring via Caesarean. Of the 28 case-control studies reporting on Caesarean delivery as an outcome, 12 studies found a significant association with maternal SARS-CoV-2, although these studies were much larger than those not finding an association (including 651,224 births as compared to 9751 births).

Preterm birth (<37 gestational weeks) in SARS-CoV-2-affected pregnancies occurred at weighted mean rate of 14%. The median prematurity rate in SARS-CoV-2 affected pregnancies was 16%, owing to four smaller studies finding very high rates of prematurity. Most larger studies reported a higher risk of preterm birth (10 studies including 648,804 births), but several smaller studies did not (10 studies including 9807 births, see figure 3). Prematurity rates in pregnancies affected by SARS-CoV-2 were not significantly different across income categories, except for rates being significantly higher in upper-middle income countries (mean 22.7%) compared to high income countries (mean 16.3%, p=0.043).

**Figure 3:**
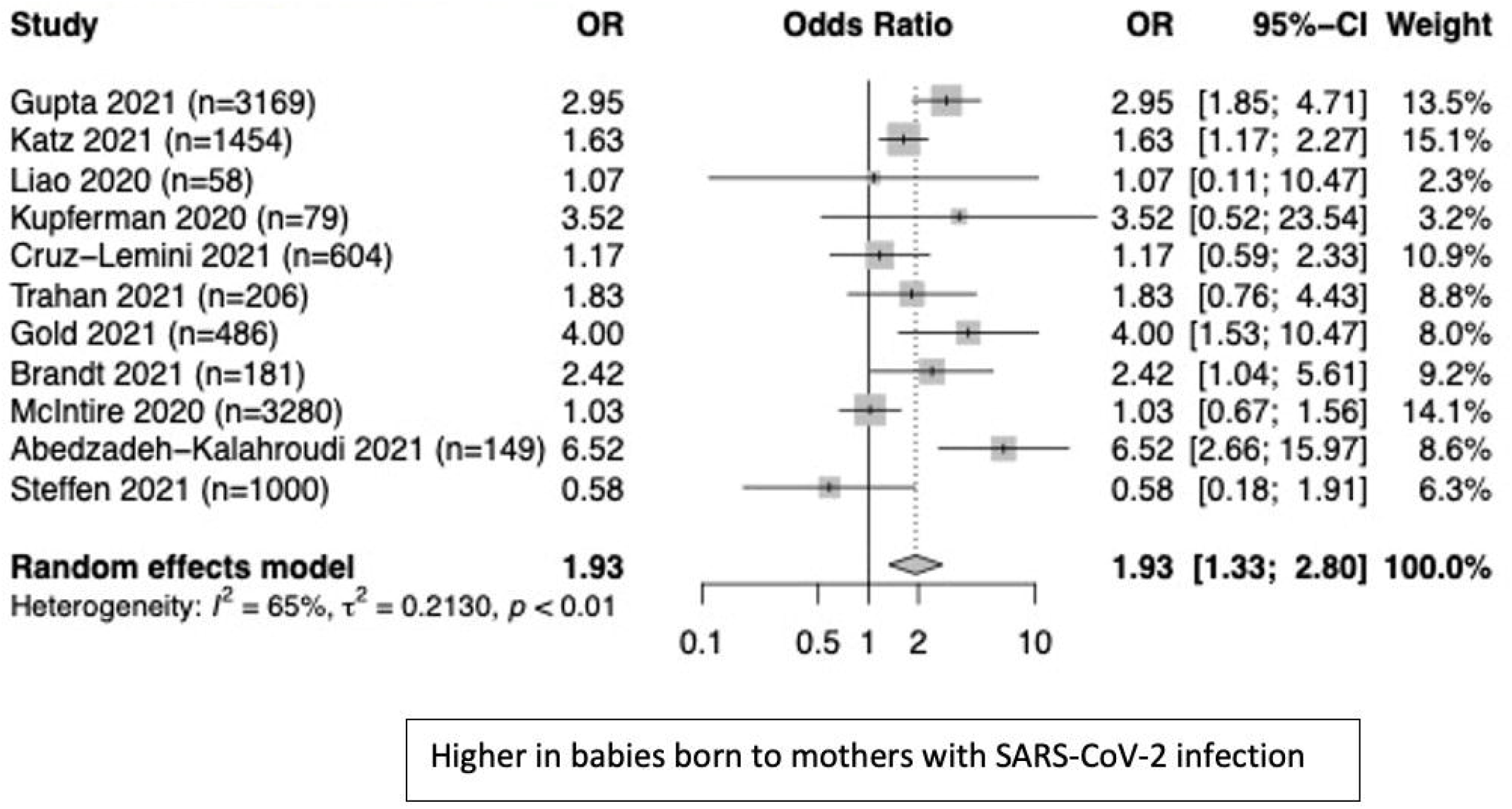
Forest plot for premature delivery

54 studies reported rates of small for gestational age births, including 753,945 infants. The range was 0-44%, and the weighted mean was 4%. 25 studies examined the rates of low birthweight. These included only 5108 infants and found a range of low birthweight rates of 0-50%, with a weighted mean of 1%.

### Breastfeeding

Breastfeeding rates among babies born to mothers with SARS-CoV-2 varied significantly across the 39 studies (96,174 infants) reporting this outcome: 0%-100% (weighted mean 12%). Of the studies reporting breastfeeding as an outcome, 28.2% reported breastfeeding status at hospital discharge, and 20.5% reported breastfeeding status at hospital discharge. The longest follow up of breastfeeding was 2 months, in 3 studies. In 7 studies it was unclear at what point breastfeeding status was recorded.

Four case-control studies including 88,567 babies examined breastfeeding by maternal SARS-CoV-2 infection status; 2 small studies (145 infants) found a significant negative association between maternal SARS-CoV-2 infection and breastfeeding[21][23], whereas 2 other studies (88,422 infants) did not find any significant association between maternal SARS-CoV-2 and breastfeeding. Among studies without a SARS-CoV-2-negative comparator group, one found that asymptomatic mothers were more likely to breastfeed than those with symptoms[24], and one found a significant difference in breastfeeding rates both in hospital and at home between those that were separated (0% in hospital, 12.2% at home) from their babies and those that were not (22.2% in hospital, 27.8% at home)[25].

### Neurodevelopmental outcomes

Two cohort studies of 339 infants examined developmental outcomes. One study found that psychomotor development was normal at 6 months in all 282 infants born following maternal SARS-CoV-2 infection during pregnancy[26]. A second study examined neurobehavioural development using the Ages and Stages questionnaire at 3 months in 57 exposed infants[27], and found that 63.6% had concerning features in the social-emotional developmental domain[27], and that abnormal development was associated with length of mother-baby separation[27]. Two studies of 191 infants found higher rates of abnormal auditory brainstem response hearing tests (44.9% vs 23.7%) and poorer otoacoustic emission test results in babies born to mothers infected with SARS-CoV-2[28][29].

### Mortality

In all studies reporting neonatal or infant mortality, there were 512 deaths reported. 10 case-control studies of 96,688 infants examined neonatal mortality, and none found a significant difference in mortality rate between neonates born to infected mothers and controls. The only study in a low income country reported no neonatal deaths[30].

## Discussion

We report the largest systematic review of neonatal and infant outcomes of babies born to women with SARS-CoV-2 in pregnancy, including 57,059 pregnancies and 45,646 babies where mothers had been infected with SARS-CoV-2 during pregnancy from 114 countries. The exclusion of case series and case reports reduced the impact of selection bias, and we excluded duplicate populations from our analysis. Building on previous studies which concentrated on timing and method of delivery [3][4], we have examined available data on neonatal morbidity which may have long-term consequences. Additionally, we included pregnancies with a maternal SARS-CoV-2 infection irrespective of whether the mother was symptomatic or asymptomatic, in contrast to earlier studies focusing on hospitalised or severely unwell mothers. Unfortunately, limited study numbers made it impossible to meta-analyse outcomes in symptomatic women compared to asymptomatic women.

As in other reviews, we found that maternal infection with SARS-CoV-2 during pregnancy is associated with higher rates of prematurity [8][31][32]. We found that prematurity rates were highest in upper-middle income countries, although they were similar in lower-middle income countries. This could be due to iatrogenic premature delivery rather than spontaneous preterm labour, but more study will be required to determine the aetiology.

We also found some evidence that maternal infection with SARS-CoV-2 is associated with increased rates of admission to the neonatal intensive care unit. The reason for this could be the increase in prematurity, as reported above, but it should be noted that some of these admissions may be for isolation purposes, an observation period, or for the care of a baby whose mother is severely unwell and unable to care for the baby herself. Additionally, in resource-limited settings, specialist neonatal intensive care may not be available – hence, this is not a generalisable marker for neonatal morbidity in all settings.

Evidence is limited and conflicting as to the association between maternal SARS-CoV-2 in pregnancy and short or long-term neonatal morbidity. The strongest evidence supports an association between maternal infection and an increased risk of respiratory disease mediated by preterm birth, but not of neurological or gastrointestinal morbidity[22]. We identified few, small studies which examined longer-term developmental outcomes; these found an apparent association between maternal SARS-CoV-2 infection and adverse outcomes in early infancy (3 months), but more studies that follow infants up over a longer time period will be needed to determine the true effect of maternal SARS-CoV-2 infection on development. It is imperative that these concerning findings are examined using standardised and validated neurodevelopmental assessments, and with the same assessment tools throughout multiple studies to allow meta-analysis. These findings also highlight the critical importance of examining neurodevelopment of offspring exposed to SARS-CoV-2 in-utero or in early life definitively through larger studies. Two small studies reported an association between maternal SARS-CoV-2 infection and offspring hearing impairment in healthy newborns without any specific risk factors for hearing impairment [28][29], further supporting the importance of following up children exposed to SARS-CoV-2 in pregnancy.

We were unable to examine the impact by trimester of maternal SARS-CoV-2 infection due to a paucity of studies examining offspring of first or second trimester infection. Other viruses such as Zika virus are known to be harmful to the developing foetus when contracted in the first or second trimester[33], so there is a reasonable suspicion that this could be true for SARS-CoV-2. Future studies should focus on examining this critical question, particularly as the virus becomes endemic.

Our study did not find clear evidence that maternal SARS-CoV-2 infection is associated with a reduction in breastfeeding. Reductions reported in some studies may relate to mother-baby separation or maternal symptoms as opposed to a direct effect of the virus: one study finding lower breastfeeding rates in cases was based in China, which recommended against breastfeeding if a lactating woman was infected with SARS-CoV-2[23]. Those finding no difference were based in Sweden[22] where there were no recommendations to restrict breastfeeding, and in the USA[34]; we were unable to verify the exact guidance used by USA-based study hospitals at the time of data collection. We chose not to report vertical transmission of SARS-CoV-2 in this review, as identified studies varied widely in the timing and type of SARS-CoV-2 testing undertaken in newborns, making a true diagnosis of vertical transmission difficult to accurately report.

Reassuringly, we did not find any evidence of an increased risk of neonatal or infant death with maternal SARS-CoV-2 infection. This is in contrast to other coronaviruses such as MERS, which has been linked with neonatal mortality rates of up to 33%[4]. However, in the studies we identified, it was difficult to determine which neonatal or infant deaths might be attributable specifically to SARS-CoV-2 infection during pregnancy. We elected not to include case reports and case series in this review, but it should be noted that cases of severe SARS-CoV-2 infections in neonates have been reported[35][36][37]. Although the incidence is likely to be low, this review does not seek to exclude severe neonatal infection with SARS-CoV-2 as a possibility.

This review identifies a crucial lack of data regarding the consequences for women in lower-income settings. Our findings suggest that some of the adverse perinatal outcomes may be more common in lower-middle and upper-middle income countries than in high-income countries, such as prematurity, but we had insufficient evidence to determine whether this trend continued into low-income countries. Birth rates are consistently higher in lower-income settings[38], and so many more pregnant women may be affected by SARS-CoV-2 infection in these regions[39] where specialist neonatal care may be limited.

Our study has several limitations. Although we identified many studies reporting perinatal outcomes, there was little information reporting neonatal morbidity in depth. Granular detail describing the indirect neonatal consequences of maternal SARS-CoV-2 infection during pregnancy remain unclear. This limitation is particularly pronounced for neurodevelopmental outcomes. With the SARS-CoV-2 declared pandemic two years ago, we hope that more information regarding these crucial outcomes will emerge soon; one trial is currently recruiting (the ASPIRE trial) which will follow up infant outcomes for 1.5 years[40], and another (the SINEPOST study) will examine development from 18 months onwards[41]. Furthermore, we found that studies varied widely on their reporting of severity of maternal disease and maternal symptoms; therefore, we were unable to study the effect of maternal symptomology on neonatal outcomes. Finally, we found limited evidence from middle-, and particularly, low-income countries, and little data regarding infections in early pregnancy. These are key research priorities to allow clinicians to adequately inform expectant families.

## Conclusion

There is a lack of evidence surrounding neonatal morbidity and longer-term outcomes for babies born to SARS-CoV-2 infected mothers. Neonatal and child health researchers should attempt to address this crucial evidence gap to adequately inform families, healthcare professionals, and public health responses.

### What is already known on this topic

SARS-CoV-2 infection in pregnancy is known to be associated with increased risk of obstetric morbidity, including need for intensive care or Caesarean delivery. Viral infection during pregnancy can be associated with adverse neurodevelopmental outcomes. It is therefore of clinical and public health significance to understand the risks of maternal SARS-CoV-2 for neonates and infants.

### What this study adds

This study summarises studies of maternal SARS-CoV-2 in pregnancy with a specific focus on neonatal outcomes. Affected pregnancies were associated with an increased risk of preterm birth, and admission to the neonatal unit. However, a significant research gap was identified in relation to individual neonatal morbidities, with limited data from lower-income settings and early pregnancy infections.

## Supporting information

PRISMA flow chart

Appendix 1 - Search terms

Appendix 2 - Results table

## Data Availability

All data produced in the present study are available upon reasonable request to the authors

## Contributorship statement

The study was designed by SS and SA with input from CG, CB and KLD. Data extraction and analysis was completed by SS and SA, with CG participating where there was disagreement over inclusion of papers. SS prepared the manuscript, which was reviewed and edited by all.

## Data sharing statement

The authors are willing to share their data upon request with any interested parties, including for future research.

## References

1 World Health Organization. Coronavirus disease (COVID-19): Pregnancy and childbirth. 2021.https://www.who.int/news-room/q-a-detail/coronavirus-disease-covid-19-pregnancy-and-childbirth (accessed 17 Sep 2021).

2 Galang RR, Chang K, Strid P, et al. Severe coronavirus infections in pregnancy: A systematic review. Obstet Gynecol 2020;136:262–72. doi:10.1097/AOG.0000000000004011

3 Diriba K, Awulachew E, Getu E. The effect of coronavirus infection (SARS-CoV-2, MERS-CoV, and SARS-CoV) during pregnancy and the possibility of vertical maternal-fetal transmission: a systematic review and meta-analysis. Eur J Med Res 2020;25:1–14. doi:10.1186/s40001-020-00439-w

4 Di Mascio D, Khalil A, Saccone G, et al. Outcome of coronavirus spectrum infections (SARS, MERS, COVID-19) during pregnancy: a systematic review and meta-analysis. Am J Obstet Gynecol MFM Published Online First: 2020. doi:10.1016/j.ajogmf.2020.100107

5 Zhang P, Heyman T, Greechan M, et al. Maternal, neonatal and placental characteristics of SARS-CoV-2 positive mothers. J Matern Neonatal Med 2021;0:1–9. doi:http://dx.doi.org/10.1080/14767058.2021.1892637

6 Chi H, Chiu N-C, Tai Y-L, et al. Clinical features of neonates born to mothers with coronavirus disease-2019: A systematic review of 105 neonates. J Microbiol Immunol Infect 2021;54:69–76. doi:https://dx.doi.org/10.1016/j.jmii.2020.07.024

7 Fenizia C, Biasin M, Cetin I, et al. Analysis of SARS-CoV-2 vertical transmission during pregnancy. Nat Commun 2020;11:5128. doi:10.1038/s41467-020-18933-4

8 Chi J, Gong W, Gao Q, et al. Clinical characteristics and outcomes of pregnant women with COVID-19 and the risk of vertical transmission: a systematic review. Arch Gynecol Obstet 2021;303:337–45. doi:http://dx.doi.org/10.1007/s00404-020-05889-5

9 Bwire GM, Njiro BJ, Mwakawanga DL, et al. Possible vertical transmission and antibodies against SARS-CoV-2 among infants born to mothers with COVID-19: A living systematic review. J Med Virol 2021;93:1361–9. doi:http://dx.doi.org/10.1002/jmv.26622

10 Gale C, Quigley MA, Placzek A, et al. Characteristics and outcomes of neonatal SARS-CoV-2 infection in the UK: a prospective national cohort study using active surveillance. Lancet Child Adolesc Heal 2021;5:113–21. doi:https://dx.doi.org/10.1016/S2352-4642(20)30342-4

11 Royal College of Obstetricians and Gynaecologists. Zika Virus Infection and Pregnancy. 2019.

12 Gale C, Statnikov Y, Jawad S, et al. Neonatal brain injuries in England: Population-based incidence derived from routinely recorded clinical data held in the National Neonatal Research Database. Arch Dis Child Fetal Neonatal Ed 2018;103:F301–6. doi:10.1136/archdischild-2017-313707

13 Webbe JWH, Duffy JMN, Afonso E, et al. Core outcomes in neonatology: development of a core outcome set for neonatal research. Arch Dis Child -Fetal Neonatal Ed 2020;105:425–31. doi:10.1136/archdischild-2019-317501

14 Ouzzani M, Hammady H, Fedorowicz Z, et al. Rayyan—a web and mobile app for systematic reviews. Syst Rev 2016;5:210. doi:10.1186/s13643-016-0384-4

15 Wells G, Shea B, O’Connell D, et al. The Newcastle-Ottawa Scale (NOS) for assessing the quality of nonrandomised studies in meta-analyses. Ottawa Hosp. Res. Inst. http://www.ohri.ca/programs/clinical_epidemiology/oxford.asp (accessed 6 Nov 2015).

16 IBM. SPSS Statistics for Macintosh. 2020.

17 R Core Team. R: A language and environment for statistical computing. 2021.

18 World Bank. World Bank Country and Lending Groups. 2020.https://datahelpdesk.worldbank.org/knowledgebase/articles/906519-world-bank-country-and-lending-groups (accessed 21 May 2015).

19 Sturrock S, Turner K, Lee-wo C, et al. The COVID19 pandemic has changed women’s experiences of pregnancy in the UK. 2021.

20 Hcini N, Maamri F, Lambert V, et al. Maternal, fetal and neonatal outcomes of large series of SARS-CoV-2 positive pregnancies in peripartum period: A single-center prospective comparative study. Eur J Obstet Gynecol Reprod Biol 2021;257:11–8. doi:http://dx.doi.org/10.1016/j.ejogrb.2020.11.068

21 Farghaly MAA, Kupferman F, Castillo F, et al. Characteristics of Newborns Born to SARS-CoV-2-Positive Mothers: A Retrospective Cohort Study. Am J Perinatol 2020;37:1310–6. doi:10.1055/s-0040-1715862

22 Norman M, Navér L, Söderling J, et al. Association of Maternal SARS-CoV-2 Infection in Pregnancy with Neonatal Outcomes. JAMA -J Am Med Assoc 2021;325:2076–86. doi:10.1001/jama.2021.5775

23 Peng S, Zhu H, Yang L, et al. A study of breastfeeding practices, SARS-CoV-2 and its antibodies in the breast milk of mothers confirmed with COVID-19. Lancet Reg Heal - West Pacific 2020;4. doi:10.1016/j.lanwpc.2020.100045

24 Abdulghani SH, Shaiba LA, Bukhari MA, et al. Consequences of SARS-CoV-2 disease on maternal, perinatal and neonatal outcomes: A retrospective observational cohort study. Clin Exp Obstet Gynecol 2021;48:353–8. doi:http://dx.doi.org/10.31083/j.ceog.2021.02.2361

25 Popofsky S, Noor A, Leavens-Maurer J, et al. Impact of Maternal Severe Acute Respiratory Syndrome Coronavirus 2 Detection on Breastfeeding Due to Infant Separation at Birth. J Pediatr 2020;226:64–70.http://www.elsevier.com/inca/publications/store/6/2/3/3/1/1/index.htt

26 Vazquez SV, Carrasco I, Perez AP, et al. Microbiological features and follow-up of neonates born to mothers with covid-19. Top Antivir Med 2021;29:237.https://120qrk11gh163n79gg1cg656-wpengine.netdna-ssl.com/wp-content/uploads/2021/03/march-2021.pdf

27 Wang Y, Chen L, Wu T, et al. Impact of Covid-19 in pregnancy on mother’s psychological status and infant’s neurobehavioral development: a longitudinal cohort study in China. BMC Med 2020;18:1–10. doi:10.1186/s12916-020-01825-1

28 Alan MA, Alan Mehmet Akif; ORCID: http://orcid.org/0000-0002-2039-8701CAO-A. Hearing screening outcomes in neonates of SARS-CoV-2 positive pregnant women. Int J Pediatr Otorhinolaryngol 2021;146:110754. doi:http://dx.doi.org/10.1016/j.ijporl.2021.110754

29 Celik T, Koca CF, Aydin S, et al. Evaluation of cochlear functions in infants exposed to SARS-CoV-2 intrauterine. Am J Otolaryngol - Head Neck Med Surg 2021;42:102982. doi:http://dx.doi.org/10.1016/j.amjoto.2021.102982

30 Hcini N, Maamri F, Picone O, et al. Maternal, fetal and neonatal outcomes of large series of SARS-CoV-2 positive pregnancies in peripartum period: a single-center prospective comparative study. Spec Issue An Updat Infect pregnancy 2021;257:11–8. doi:http://dx.doi.org/10.1016/j.ejogrb.2020.11.068

31 Khalil A, Kalafat E, Benlioglu C, et al. SARS-CoV-2 infection in pregnancy: A systematic review and meta-analysis of clinical features and pregnancy outcomes. EClinicalMedicine Published Online First: 2020. doi:10.1016/j.eclinm.2020.100446

32 Toro F Di, Gjoka M, Lorenzo G Di, et al. Since January 2020 Elsevier has created a COVID-19 resource centre with free information in English and Mandarin on the novel coronavirus COVID-19. The COVID-19 resource centre is hosted on Elsevier Connect, the company’s public news and information 2020.

33 Souza JP, Méio MDBB, de Andrade LM, et al. Adverse fetal and neonatal outcomes in pregnancies with confirmed zika virus infection in Rio de Janeiro, Brazil: A cohort study. PLoS Negl Trop Dis 2021;15:1– 13. doi:10.1371/journal.pntd.0008893

34 Flaherman V, Afshar Y, Boscardin W, et al. Infant Outcomes Following Maternal Infection With Severe Acute Respiratory Syndrome Coronavirus 2 (SARS-CoV-2): First Report From the Pregnancy Coronavirus Outcomes Registry (PRIORITY) Study. 2020.

35 Shaiba LA, Hadid A, Altirkawi KA, et al. Case Report: Neonatal Multi-System Inflammatory Syndrome Associated With SARS-CoV-2 Exposure in Two Cases From Saudi Arabia. Front Pediatr 2021;9:1–8. doi:10.3389/fped.2021.652857

36 Sagheb S, Lamsehchi A, Jafary M, et al. Two seriously ill neonates born to mothers with COVID-19 pneumonia-a case report. Ital J Pediatr 2020;46:137.

37 Hinojosa-Velasco A, de Oca PVB-M, García-Sosa LE, et al. A case report of newborn infant with severe COVID-19 in Mexico: Detection of SARS-CoV-2 in human breast milk and stool. Int J Infect Dis 2020;100:21–4. doi:10.1016/j.ijid.2020.08.055

38 World Bank. Data Bank. 2021.https://databank.worldbank.org/reports.aspx?source=2&series=SP.DYN.IMRT.IN&country=LIC,HI,MICC (accessed 15 Jun 2021).

39 Our World in Data. Total confirmed COVID-19 cases. https://ourworldindata.org/grapher/covid-cases-income (accessed 15 Jun 2021).

40 Huddleston H, Jaswa E, Gaw S, et al. ASPIRElll: A ssessing the S afety of P regnancy I n the Co R onavirus Pand E mic Our Team Team. 2021;:2021.https://aspire.ucsf.edu (accessed 4 Aug 2021).

41 Action Medical Research For Children. COVID-19 – understanding the impact of exposure to SARS-CoV-2 early in life on a child’s brain development and mental health. 2021.https://action.org.uk/research/covid-19-understanding-impact-exposure-sars-cov-2-early-life-childs-brain-development-and (accessed 4 Aug 2021).

