## Supplementary material for "Neonatal outcomes and indirect consequences following maternal SARS-CoV-2 infection in pregnancy: A systematic review": PRISMA flow chart

Figure 1: Study selection flow chart

Records identified from:

Databases (n = 5352)

Registers (n = 22)

Records removed *before screening*:

Duplicate records removed (n = 2140)

Reports assessed for eligibility

(n = 3234)

Reports excluded:

Study design (review paper, case report or case series) (n = 1371)

Study population (not pregnant women, not SARS-CoV-2 infection) (n = 704)

Study outcome (n = 419)

Duplicate records or data (n = 236)

Publication date pre-1^st^ January 2020	(n=300)

Studies included in review

(n = 204)

Cohort studies = 167

Case-control studies = 37

**Identification of studies via databases and registers**

**Identification**

**Screening**

**Included**
