## Appendix 1 - Search terms for "Neonatal outcomes and indirect consequences following maternal SARS-CoV-2 infection in pregnancy: A systematic review"

Appendix 1 – Search strategy

Embase search via Ovid, last updated 28/07/2021.

| # | Searches | Results |
|---|---|---|
| 1 | Pregnan* | 723025 |
| 2 | Third trimester pregnancy/ or first trimester pregnancy/ or "parameters concerning the fetus, newborn and pregnancy"/ or pregnancy/ or pregnancy complication/ or high risk pregnancy/ or pregnancy disorder/ or second trimester pregnancy/ or pregnancy outcome/ | 498419 |
| 3 | Antenat* | 51397 |
| 4 | Gestat* | 297546 |
| 5 | Matern* | 379690 |
| 6 | Maternity ward/ or maternity care/ | 21247 |
| 7 | Breastfeed* | 35731 |
| 8 | Breast feeding/ or mother/ or breast milk/ | 140234 |
| 9 | Lactat* | 227694 |
| 10 | Lactation/ | 39318 |
| 11 | 1 or 2 or 3 or 4 or 5 or 6 or 7 or 8 or 9 or 10 | 1220052 |
| 12 | COVID* | 147977 |
| 13 | Sars-cov-2 | 52773 |
| 14 | Coronavirus | 166814 |
| 15 | Severe acute respiratory syndrome coronavirus 2/ or coronavirus disease 2019/ or coronavirus infection/ | 146237 |
| 16 | 12 or 13 or 14 or 15 | 187862 |
| 17 | Stillb* | 23397 |
| 18 | Stillbirth/ or congenital malformation/ | 98283 |
| 19 | Intrauterine adj demise | 239 |
| 20 | Intrauterine adj death | 1647 |
| 21 | Pregnancy adj loss | 10662 |
| 22 | Miscarr* | 25084 |
| 23 | Spontaneous adj abortion | 39613 |
| 24 | Spontaneous abortion/ | 38012 |
| 25 | Fetus death/ | 15764 |
| 26 | 17 or 18 or 19 or 20 or 21 or 22 or 23 or 24 or 25 | 158662 |
| 27 | Neonat* | 303927 |
| 28 | Infan* | 701133 |
| 29 | Newborn/ | 366307 |
| 30 | Infant/ or hospitalized infant/ or infant disease/ or infant mortality/ or high risk infant/ or infant care/ | 477647 |
| 31 | 27 or 28 or 29 or 30 | 995198 |
| 32 | Infect* | 2546071 |
| 33 | Infection/ or infection complication/ or infection risk/ or intrauterine infection/ or perinatal infection/ | 378561 |

| | | |
|---|---|---|
| 34 | Sepsis | 201867 |
| 35 | Newborn sepsis/ or sepsis/ | 157457 |
| 36 | Nicu or neonatal adj admission | 2739 |
| 37 | Newborn intensive care/ or neonatal intensive care unit/ | 37168 |
| 38 | Premat* | 263202 |
| 39 | Prematurity/ or premature labour/ | 130336 |
| 40 | Small adj for adj gestational adj age or sga | 20340 |
| 41 | Small for date infant/ | 15970 |
| 42 | Low adj birthweight | 7252 |
| 43 | Low birth weight/ | 31063 |
| 44 | Operative adj delivery | 1711 |
| 45 | Instrumental delivery/ | 3855 |
| 46 | Caesarean or c-section or cesarean | 107540 |
| 47 | Cesarean section/ | 92477 |
| 48 | Instrumental delivery or ventouse or forceps or vacuum extraction | 27358 |
| 49 | Vacuum extraction/ or forceps delivery/ or forceps/ | 13907 |
| 50 | Ventilat* | 291355 |
| 51 | Respiratory adj distress | 100158 |
| 52 | CPAP or BiPAP | 18918 |
| 53 | Ventilated patient/ or artificial ventilation/ | 137133 |
| 54 | Continuous positive airway pressure/ or bilevel positive airway pressure | 3083 |
| 55 | Respiratory distress/ | 32246 |
| 56 | Seizure or convulsion or fit | 361565 |
| 57 | Seizure/ or ”seizure, epilepsy or convulsion”/ | 133064 |
| 58 | Hypoxic ischaemic encephalopathy or hie | 12140 |
| 59 | Hypoxic ischaemic encephalopathy/ | 8490 |
| 60 | Therapeutic cooling or induced hypothermia | 171749 |
| 61 | Brain ischemia/ or induced hypothermia/ or cooling/ | 171749 |
| 62 | MRI or magnetic resonance imaging or eeg or electroencephalogram or cruss or cranial ultrasound or brain scan or brain imaging | 1108432 |
| 63 | Neuroimaging/ or nuclear magnetic resonance imaging/ or functional magnetic resonance imaging/ or electroencephalogram/ | 1011212 |
| 64 | Hypoton* or hyperton* or abnormal tone or spasticity or cerebral palsy or neurological disease or neurological abnormality | 118366 |
| 65 | Spasticity/ or cerebral palsy/ or muscle hypertonia/ or neurologic disease/ | 176148 |
| 66 | Cognitive ability or learning difficulty or learning disability or developmental delay | 36605 |
| 67 | Learning disorder/ or developmental delay/ or developmental disorder/ or mental deficiency/ | 93582 |

| | | |
|---|---|---|
| 68 | Gastrointestinal disease or nec or necrotising enterocolitis or necrotizing enterocolitis | 104188 |
| 69 | Gastrointesintal disease/ or necrotizing enterocolitis/ | 95690 |
| 70 | Visual impairment or visually impaired or blind* | 494101 |
| 71 | Visual impairment/ or blindness/ | 73184 |
| 72 | Hearing impair* or deaf* | 87073 |
| 73 | Hearing impairment/ | 46891 |
| 74 | Quality of life or "quality of life"/ | 637206 |
| 75 | Vertical transmi* | 17715 |
| 76 | Vertical transmission/ | 14908 |
| 77 | 32-76, combined with OR | 5907734 |
| 78 | 77 and 31 | 459481 |
| 79 | 78 or 26 | 593511 |
| 80 | 11 and 16 and 79 | 1824 |

Medline search via Ovid, last updated 28/07/2021.

| # | Searches | Results |
|---|---|---|
| 1 | Pregnan* | 1047802 |
| 2 | Pregnancy trimester, third/ or pregnancy trimester, second/ or pregnancy trimester, first/ or pregnancy/ or pregnancy, high-risk/ or pregnancy outcome/ or pregnancy complications/ | 911420 |
| 3 | Antenat* | 41206 |
| 4 | Prenatal care/ | 29574 |
| 5 | Gestat* | 261469 |
| 6 | Matern* | 359413 |
| 7 | Perinatal care/ | 4983 |
| 8 | Breastfeed* | 30017 |
| 9 | Breast feeding/ or mothers/ | 81408 |
| 10 | Lactat* | 224527 |
| 11 | Lactation/ | 44082 |
| 12 | 1 or 2 or 3 or 4 or 5 or 6 or 7 or 8 or 9 or 10 or 11 | 1454976 |
| 13 | COVID* | 156147 |
| 14 | Sars-cov-2 | 99388 |
| 15 | Coronavirus | 98845 |
| 16 | Covid-19/ or sars-cov-2/ or coronavirus/ | 97776 |
| 17 | 13 or 14 or 15 or 16 | 175760 |
| 18 | Stillb* | 17756 |
| 19 | Stillbirth/ | 5353 |
| 20 | Intrauterine adj demise | 142 |
| 21 | Intrauterine adj death | 1477 |
| 22 | Pregnancy adj loss | 6997 |
| 23 | Miscarr* | 15892 |
| 24 | Spontaneous adj abortion | 7719 |

| | | |
|---|---|---|
| 25 | Spontaneous abortion/ | 20453 |
| 26 | Fetal death/ or congenital abnormalities/ | 57641 |
| 27 | 18 or 19 or 20 or 21 or 22 or 23 or 24 or 25 or 26 | 106523 |
| 28 | Neonat* | 309101 |
| 29 | Infan* | 1335539 |
| 30 | Infant, newborn/ | 626610 |
| 31 | Infant/ or infant care/ or infant mortality/ or infant death/ or infant health/ | 841541 |
| 32 | 28 or 29 or 30 or 31 | 1450833 |
| 33 | Infect* | 2447138 |
| 34 | Infections/ | 39946 |
| 35 | Sepsis | 133814 |
| 36 | Neonatal sepsis/ or sepsis/ | 63823 |
| 37 | Nicu or neonatal admission | 11656 |
| 38 | Intensive care units, neonatal/ | 15858 |
| 39 | Premat* | 223778 |
| 40 | Infant, premature/ or gestational age/ or premature birth/ | 139364 |
| 41 | Small for gestational age or sga | 16965 |
| 42 | Infant, small for gestational age/ or birth weight/ | 46772 |
| 43 | Low adj birthweight | 7841 |
| 44 | Infant, low birth weight/ | 19191 |
| 45 | Operative adj delivery | 1256 |
| 46 | Extraction, obstetrical/ | 2534 |
| 47 | Caesarean or c-section or cesarean | 78432 |
| 48 | Cesarean section/ or delivery, obstetric/ | 72546 |
| 49 | Instrumental delivery or ventouse or forceps or vacuum extraction | 14655 |
| 50 | Vacuum extraction, obstetrical/ or obstetrical forceps/ | 2720 |
| 51 | Ventilat* | 201282 |
| 52 | Respiratory adj distress | 61778 |
| 53 | CPAP or BiPAP or Continuous positive airway pressure or bilevel positive airway pressure | 16071 |
| 54 | Ventilation/ or respiration, artificial/ | 58137 |
| 55 | Continuous positive airway pressure/ or positive-pressure respiration/ or noninvasive ventilation/ | 27370 |
| 56 | Respiratory distress syndrome/ | 21490 |
| 57 | Seizure or convulsion or fit | 208598 |
| 58 | Seizures/ | 56493 |
| 59 | Hypoxic ischaemic encephalopathy or hypoxic ischemic encephalopathy or hie | 5083 |
| 60 | Hypoxia-ischaemia, brain/ or asphyxia neonatorum | 13249 |
| 61 | Therapeutic cooling or induced hypothermia | 2598 |
| 62 | Brain ischemia/ or hypothermia, induced/ | 75818 |

| | | |
|---|---|---|
| 63 | MRI or magnetic resonance imaging or eeg or electroencephalogram or cruss or cranial ultrasound or brain scan or brain imaging | 707043 |
| 64 | Neuroimaging/ or magnetic resonance imaging/ or electroencephalogram/ | 575390 |
| 65 | Hypoton* or hyperton* or abnormal tone or spasticity or cerebral palsy or neurological disease or neurological abnormality | 103230 |
| 66 | Muscle spasticity/ or cerebral palsy/ or muscle hypertonia/ or muscle hypotonia/ or nervous system diseases/ | 78506 |
| 67 | Cognitive ability or learning difficulty or learning disability or developmental delay | 23602 |
| | Learning disabilities/ or developmental disabilities/ or intellectual disability/ | 87101 |
| 69 | Gastrointestinal disease or nec or necrotising enterocolitis or necrotizing enterocolitis | 15271 |
| 70 | Gastrointestinal diseases/ or enterocolitis, necrotizing/ | 43975 |
| 71 | Visual impairment or visually impaired or blind* | 395483 |
| 72 | Vision disorders/ | 28486 |
| 73 | Hearing impair* or deaf* | 62891 |
| 74 | Hearing loss/ or deafness/ or persons with hearing impairments/ | 43064 |
| 75 | Quality of life or “quality of life”/ | 380511 |
| 76 | Vertical transmi* | 7322 |
| 77 | Infectious disease transmission, vertical/ | 17257 |
| 78 | 33-77, combined with OR | 5028529 |
| 79 | 78 and 32 | 548597 |
| 80 | 79 or 27 | 636683 |
| 81 | 12 and 17 and 80 | 1143 |

Global health search via Ovid, last updated 28/07/2021

| # | Searches | Results |
|---|---|---|
| 1 | Pregnan* | 126717 |
| 2 | Pregnancy trimester, third/ or pregnancy trimester, second/ or pregnancy trimester, first/ or pregnancy/ or pregnancy, high-risk/ or pregnancy outcome/ or pregnancy complications/ | 99638 |
| 3 | Antenat* | 17826 |
| 4 | Prenatal care/ | 3506 |
| 5 | Gestat* | 112821 |
| 6 | Matern* | 86406 |
| 7 | Breastfeed* | 25392 |
| 8 | Breast feeding/ or mothers/ | 51651 |

| | | |
|---|---|---|
| 9 | Lactat* | 50673 |
| 10 | Lactation/ | 12941 |
| 11 | 1 or 2 or 3 or 4 or 5 or 6 or 7 or 8 or 9 or 10 | 229179 |
| 12 | COVID* | 36404 |
| 13 | Sars-cov-2 | 15628 |
| 14 | Coronavirus | 44855 |
| 15 | Severe acute respiratory syndrome coronavirus/ | 5256 |
| 16 | 12 or 13 or 14 or 15 | 46013 |
| 18 | Stillb* | 4753 |
| 19 | Stillbirth/ | 4006 |
| 20 | Intrauterine adj demise | 9 |
| 21 | Intrauterine adj death | 139 |
| 22 | Pregnancy adj loss | 923 |
| 23 | Miscarr* | 3987 |
| 24 | Spontaneous adj abortion | 3435 |
| 25 | Spontaneous abortion/ | 2942 |
| 26 | Fetal death/ or congenital abnormalities/ | 15922 |
| 27 | 18 or 19 or 20 or 21 or 22 or 23 or 24 or 25 or 26 | 21539 |
| 28 | Neonat* | 59142 |
| 29 | Infan* | 156485 |
| 30 | Neonates/ or infants/ | 122421 |
| 31 | Infant care/ or infant mortality/ or infant death/ or infant health/ | 5195 |
| 32 | 28 or 29 or 30 or 31 | 176742 |
| 33 | Infect* | 1187975 |
| 34 | Infections/ | 488566 |
| 35 | Sepsis | 25827 |
| 36 | Sepsis/ | 18064 |
| 37 | Nicu or neonatal admission | 2676 |
| 38 | Premat* | 32208 |
| 39 | Premature infants/ or prematurity/ | 20675 |
| 40 | Small for gestational age or sga | 4240 |
| 41 | Infant, small for gestational age/ or birth weight/ | 13532 |
| 42 | Low adj birthweight | 3185 |
| 43 | Low birth weight infants/ | 8798 |
| 44 | Operative adj delivery | 117 |
| 45 | Caesarean or c-section or cesarean | 9334 |
| 46 | Caesarean section/ | 5734 |
| 47 | Instrumental delivery or ventouse or forceps or vacuum extraction | 863 |
| 48 | Parturition complications/ | 1384 |
| 49 | Ventilat* | 20633 |
| 50 | Respiratory adj distress | 6548 |
| 51 | CPAP or BiPAP or Continuous positive airway pressure or bilevel positive airway pressure | 701 |
| 52 | Ventilation/ or artificial respiration/ | 4243 |

| | | |
|---|---|---|
| 53 | Acute respiratory distress syndrome/ | 1453 |
| 54 | Seizure or convulsion or fit | 27459 |
| 55 | Seizures/ | 3152 |
| 56 | Hypoxic ischaemic encephalopathy or hypoxic ischemic encephalopathy or hie | 456 |
| 57 | Therapeutic cooling or induced hypothermia | 157 |
| 58 | MRI or magnetic resonance imaging or eeg or electroencephalogram or cruss or cranial ultrasound or brain scan or brain imaging | 15860 |
| 59 | Magnetic resonance imaging/ or electroencephalogram/ | 6065 |
| 60 | Hypoton* or hyperton* or abnormal tone or spasticity or cerebral palsy or neurological disease or neurological abnormality | 5668 |
| 61 | Cerebral palsy/ or nervous system diseases/ | 18651 |
| 62 | Cognitive ability or learning difficulty or learning disability or developmental delay | 1816 |
| 63 | Learning disabilities/ or mental disorders/ or people with mental disabilities/ | 49942 |
| 64 | Gastrointestinal disease or nec or necrotising enterocolitis or necrotizing enterocolitis | 3697 |
| 65 | Gastrointestinal diseases/ | 10201 |
| 66 | Visual impairment or visually impaired or blind* | 48935 |
| 67 | Vision disorders/ | 3331 |
| 68 | Hearing impair* or deaf* | 5024 |
| 69 | Hearing impairment/ or deafness/ or people with hearing impairment/ | 4259 |
| 70 | Quality of life or “quality of life”/ | 35856 |
| 71 | Vertical transmi* | 5345 |
| 72 | Vertical transmission/ | 3839 |
| 73 | 33-72, combined with OR | 1392378 |
| 74 | 72 and 32 | 93930 |
| 75 | 74 or 27 | 110446 |
| 76 | 75 and 16 and 11 | 570 |

LILACS

| | | |
|---|---|---|
| 1 | SARS-CoV-2 or COVID or Coronavirus | 8903 |
