## Appendix 2 - Results table for "Neonatal outcomes and indirect consequences following maternal SARS-CoV-2 infection in pregnancy: A systematic review"

| | Title | Year | First author | Country | Start date | End date | Type of study | Number of mothers total | Number of case mothers | Number of babies total | Number of case babies | Population | Exposure |
|---|---|---|---|---|---|---|---|---|---|---|---|---|---|
| 1 | Vertical transmission and kidney damage in newborns whose mothers had coronavirus disease 2019 during pregnancy | 2020 | He, Zheng | China | 01/01/2020 | 01/04/2020 | Cohort | 22 | 22 | 22 | 22 | born to mothers with COVID-19 at one centre | COVID-19 diagnosed on clinical criteria |
| 2 | Increased C-sections and preterm births in SARS-CoV-2 infection during pregnancy | 2021 | Carrasco, I | Spain | 15/03/2020 | 31/07/2020 | Cohort | 105 | 105 | 107 | 107 | All pregnant women | PCR-confirmed COVID-19 |
| 3 | Microbiological features and follow-up of neonates born to mothers with covid-19 | 2021 | Vazquez, S | Spain | 15/03/2020 | 30/11/2020 | Cohort | 115 | 115 | 282 | 282 | neonates, 130 in first wave and 152 in second | confirmed COVID-19 infection, unclear of method of confirmation |
| 4 | COVID-19 infection in high-risk south african pregnancies with and without HIV | 2021 | De Waard, | South Africa | 01/05/2020 | 31/07/2020 | Cohort | 100 | 100 | 91 | 91 | All pregnant women | confirmed COVID-19 infection, unclear of method of confirmation |
| 5 | The impact of the novel coronavirus infection COVID-19 on the mother-placenta-fetus system | 2021 | Nizyaeva, N | Russia | 01/03/2020 | 01/05/2020 | Cohort | 66 | 66 | 42 | 42 | All pregnant women. Multiple pregnancies excluded. | COVID-19 and control group with negative PCR test and no signs of COVID-19 |
| 6 | Clinical outcomes of maternal and neonate with COVID-19 infection - Multicenter study in Saudi Arabia | 2021 | Al-Matary, | Saudi Arabia | 01/03/2020 | 01/11/2020 | Cohort | 288 | 288 | 200 | 200 | All pregnant women | PCR-confirmed COVID-19 |

| | | | | | | | | | | | | | |
|---|---|---|---|---|---|---|---|---|---|---|---|---|---|
| 7 | Characteristics, clinical and laboratory data and outcomes of pregnant women with confirmed SARS-CoV-2 infection admitted to Al-Zahra tertiary referral maternity center in Iran: a case series of 24 patients | 2020 | Vaezi, Mar | Iran | 10/03/2020 | 15/04/2020 | Cohort | 24 | 24 | 11 | 11 | All pregnant women | Laboratory-confirmed COVID-19 infection, unclear of method of confirmation |
| 8 | Outcomes of newborns to mothers with COVID-19 | 2021 | Ghema, K. | Morocco | 01/01/2020 | 01/12/2020 | Cohort | 30 | 30 | 30 | 30 | All neonates admitted to NICU whose mothers had COVID-19 | |
| 9 | SARS-CoV-2 prevalence and maternal-perinatal outcomes among pregnant women admitted for delivery: Experience from COVID-19-dedicated maternity hospital in Jammu, Jammu and Kashmir (India) | 2021 | Gupta, Pur | India | 01/09/2020 | 30/11/2020 | Case-control | 3165 | 108 | 3165 | 108 | All pregnant women delivering during study period | PCR-confirmed COVID-19 |
| 10 | Prevalence, clinical features, and outcomes of SARS-CoV-2 infection in pregnant women with or without mild/moderate symptoms: Results from universal screening in a tertiary care center in Mexico City, Mexico | 2021 | Cardona-P | Mexico | 22/04/2020 | 25/05/2020 | Case-control | 250 | 70 | 219 | 39 | All pregnant women who were asymptomatic or with mild/moderate COVID-19 symptoms | PCR-confirmed COVID-19 |
| 11 | Course of Covid 19 and fetomaternal outcome at a tertiary care hospital | 2021 | Furrukh, R | Pakistan | 08/04/2020 | 07/07/2020 | Cohort | 47 | 47 | 24 | 24 | All pregnant women | PCR-confirmed COVID-19 |

| | | | | | | | | | | | | | |
|---|---|---|---|---|---|---|---|---|---|---|---|---|---|
| 12 | Clinical and obstetric characteristics of pregnant women with Covid-19: A case series study on 26 patients | 2021 | Abedzadeh | Iran | 01/03/2020 | 01/05/2020 | Cohort | 56 | 56 | 55 | 55 | All pregnant women | PCR or radiological diagnosis of COVID-19 |
| 13 | Fetal and perinatal outcome following first and second trimester covid-19 infection: Evidence from a prospective cohort study | 2021 | Rosen, Had | Israel | 01/03/2020 | 01/02/2021 | Cohort | 55 | 55 | 29 | 29 | All pregnant women | PCR-confirmed COVID-19 before 26 weeks gestation |
| 14 | Hearing screening outcomes in neonates of SARS-CoV-2 positive pregnant women | 2021 | Alan, Mehr | Turkey | 01/04/2020 | 01/12/2020 | Cohort | 141 | 141 | 118 | 118 | born to mothers with COVID-19, multiple gestation and risk factors for hearing loss excluded | PCR-confirmed COVID-19 |
| 15 | Perinatal outcomes of pregnancies resulting from assisted reproduction technology in SARS-CoV-2-infected women: a prospective observational study | 2021 | Engels Calv | Spain | 26/02/2020 | 05/11/2020 | Cohort | 1347 | 1347 | 1347 | 1347 | All pregnant women | PCR-confirmed COVID-19 |
| 16 | The incidence, characteristics and outcomes of pregnant women hospitalized with symptomatic and asymptomatic SARS-CoV-2 infection in the UK from March to September 2020: A national cohort study using the UK Obstetric Surveillance System (UKOSS) | 2021 | Bunch, Kat | UK | 01/03/2020 | 31/08/2020 | Cohort | 1148 | 1148 | 1019 | 1019 | All hospitalised pregnant women | PCR-confirmed COVID-19 |

| | | | | | | | | | | | | | |
|---|---|---|---|---|---|---|---|---|---|---|---|---|---|
| 17 | Neonatal SARS-CoV-2 infections in breastfeeding mothers | 2021 | Shlomai, N | Israel | 01/03/2020 | 01/05/2020 | Cohort | 53 | 53 | 55 | 55 | All pregnant women delivering during study period | PCR-confirmed COVID-19 |
| 18 | COVID-19 in pregnancy-characteristics and outcomes of pregnant women admitted to hospital because of SARS-CoV-2 infection in the Nordic countries | 2021 | Juliusson, I | Denmark, Fir | 01/03/2020 | 30/06/2020 | Cohort | 56 | 56 | 51 | 51 | All pregnant women admitted to hospital for at least 24h | PCR-confirmed SARS-CoV-2 in 14 days before admission |
| 19 | Consequences of SARS-CoV-2 disease on maternal, perinatal and neonatal outcomes: A retrospective observational cohort study | 2021 | Abdulghan | Saudi Arabia | 01/03/2020 | 31/05/2020 | Cohort | 62 | 62 | 63 | 63 | All pregnant women | PCR-confirmed COVID-19 |
| 20 | Pre and post-natal epidemiological and clinical features of neonates born from mothers infected with COVID-19 and 14-day follow-up post discharge in Lima, Peru | 2021 | Lizama, Olg | Peru | 15/03/2020 | 30/06/2020 | Cohort | 206 | 206 | 206 | 206 | All pregnant women | Laboratory-confirmed COVID-19 infection, unclear of method of confirmation |
| 21 | Childbirth care among sars-cov-2 positive women in Italy | 2021 | Donati, Ser | Italy | 25/02/2020 | 31/07/2020 | Cohort | 525 | 525 | 538 | 538 | women; until end of March 2020 only symptomatic and contacts screened, | PCR-confirmed COVID-19 |

| | | | | | | | | | | | | | |
|---|---|---|---|---|---|---|---|---|---|---|---|---|---|
| 22 | Association of Maternal SARS-CoV-2 Infection in Pregnancy with Neonatal Outcomes | 2021 | Naver, Lars | Sweden | 11/03/2020 | 31/01/2021 | Case-control | 84719 | 2286 | 88159 | 2323 | All live-born infants, malformations excluded except PDA | PCR-confirmed COVID-19; from June 2020 all women tested vs just symptomatic women |
| 23 | Maternal and Neonatal Morbidity and Mortality among Pregnant Women with and without COVID-19 Infection: The INTERCOVID Multinational Cohort Study | 2021 | Villar, Jose | International | 02/03/2020 | 02/10/2020 | Case-control | 2130 | 706 | 2130 | 706 | All pregnant women | PCR, radiological or clinical criteria based diganosis of COVID-19 |
| 24 | Retrospective Analysis of Clinical Characteristics and Neonatal Outcomes of Pregnant Women with SARS-COV-2 Infection | 2021 | Chen, Yu a | China | 01/02/2020 | 30/03/2020 | Cohort | 8 | 8 | 9 | 9 | All pregnant women | COVID-19 diagnosed on clinical criteria |
| 25 | Mother-Infant Dyads with COVID-19 at an Urban, Safety-Net Hospital: Clinical Manifestations and Birth Outcomes | 2021 | Sabharwal, | USA | 31/03/2020 | 06/08/2020 | Cohort | 75 | 75 | 75 | 75 | All symptomatic pregnant women delivering during study period | PCR-confirmed COVID-19 |
| 26 | Effect of SARS-CoV-2 Infection on Pregnancy Outcomes in an Inner-City Black Patient Population | 2021 | Andrusier, | USA | 10/04/2020 | 10/06/2020 | Case-control | 335 | 56 | 335 | 56 | All pregnant women delivering during study period | PCR-confirmed COVID-19 |
| 27 | Short-term developmental outcomes in neonates born to mothers with COVID-19 from Wuhan, China | 2021 | Zeng, Ling- | China | 01/02/2020 | 15/05/2020 | Cohort | 68 | 68 | 72 | 72 | All neonates born to women with COVID-19 | COVID-19 diagnosed on clinical criteria |

| | | | | | | | | | | | | | |
|---|---|---|---|---|---|---|---|---|---|---|---|---|---|
| 28 | The Society for Obstetric Anesthesia and Perinatology (SOAP) COVID-19 Registry: An analysis of outcomes among pregnant women delivering during the initial SARS-CoV-2 outbreak in the United States | 2021 | Katz, Danie | USA | 19/03/2020 | 31/05/2020 | Case-control | 1454 | 490 | 1454 | 490 | All pregnant women | PCR-confirmed COVID-19 within 14 days of delivery date |
| 29 | Experience of covid-19 infections in neonates in tertiary care centre in North Karnataka, India: A prospective cohort study | 2021 | Charki, Sid | India | 01/05/2020 | 01/10/2020 | Cohort | 26 | 26 | 28 | 28 | All neonates born to women with COVID-19 | PCR-confirmed COVID-19 |
| 30 | Outcomes of Neonates Born to Mothers with Severe Acute Respiratory Syndrome Coronavirus 2 Infection at a Large Medical Center in New York City | 2021 | Walzer, La | USA | 13/03/2020 | 24/04/2020 | Cohort | 100 | 100 | 101 | 101 | All neonates born to women with COVID-19 | PCR-confirmed COVID-19 |
| 31 | [Management of labour, puerperium, and lactation in SARS-CoV-2 positive women. Multicentric study in the Valencian Community]. | 2021 | Vila-Cande | Spain | 01/03/2020 | 30/06/2020 | Cohort | 13 | 13 | 13 | 13 | All pregnant women who delivered | PCR-confirmed COVID-19 |
| 32 | Impact of evolving practices on SARS-CoV-2 positive mothers and their newborns in the largest public healthcare system in America. | 2021 | Malhotra, | USA | 01/03/2020 | 09/05/2020 | Cohort | 286 | 286 | 290 | 290 | All pregnant women who delivered | PCR-confirmed COVID-19 |
| 33 | Characteristics and Pregnancy Outcomes of Asymptomatic and Symptomatic Women with COVID-19: Lessons from Hospitals in Wuhan. | 2021 | Luo, Qingq | China | 30/01/2020 | 15/04/2020 | Cohort | 41 | 41 | 42 | 42 | All pregnant patients admitted | PCR or radiological diagnosis of COVID-19 |

| | | | | | | | | | | | | | |
|---|---|---|---|---|---|---|---|---|---|---|---|---|---|
| 34 | Association of Maternal Perinatal SARS-CoV-2 Infection With Neonatal Outcomes During the COVID-19 Pandemic in Massachusetts. | 2021 | Angelidou, | USA | 01/03/2020 | 31/07/2020 | Cohort | 250 | 250 | 255 | 255 | infant dyads whose delivery and discharge fell within the study | PCR-confirmed COVID-19 |
| 35 | SARS-COV-2 infection in pregnant women and newborns in a Spanish cohort (GESNEO-COVID) during the first wave. | 2021 | Carrasco, I | Spain | 15/03/2020 | 31/07/2020 | Cohort | 105 | 105 | 107 | 107 | All pregnant women delivering during study period | PCR, serological or clinical criteria based diagnosis of COVID-19 |
| 36 | Pregnancy and perinatal outcomes of women with coronavirus disease (COVID-19) pneumonia: a preliminary analysis. (case series?) | 2020 | Liu DeHan | China | 20/01/2020 | 10/02/2020 | Cohort | 11 | 11 | 11 | 11 | All pregnant women | PCR-confirmed COVID-19 |
| 37 | Characteristics and outcomes of pregnant women admitted to hospital with confirmed SARS-CoV-2 infection in UK: national population based cohort study. | 2020 | Knight, M. | U.K | 01/03/2020 | 14/04/2020 | Cohort | 427 | 427 | 259 | 259 | All pregnant women | PCR-confirmed COVID-19 |
| 38 | Clinical features and outcomes of pregnant women suspected of coronavirus disease 2019. | 2020 | Yang Hui a | China | 20/01/2020 | 05/03/2020 | Case-control | 55 | 13 | 57 | 13 | All pregnant women delivering during study period | PCR-confirmed COVID-19 |
| 39 | Clinical features and obstetric and neonatal outcomes of pregnant patients with COVID-19 in Wuhan, China: a retrospective, single-centre, descriptive study. | 2020 | Yu Nan and | China | 01/01/2020 | 08/02/2020 | Cohort | 7 | 7 | 7 | 7 | All pregnant women | PCR-confirmed COVID-19 |

| | | | | | | | | | | | | | |
|---|---|---|---|---|---|---|---|---|---|---|---|---|---|
| 40 | Covid-19 in pregnant women: General data from a French National Survey. | 2020 | Cohen, Jon | France | Not stated | | Cohort | 88 | 88 | 14 | 14 | All pregnant women | PCR, serological or radiological diagnosis of COVID-19 |
| 41 | Association Between Mode of Delivery Among Pregnant Women With COVID-19 and Maternal and Neonatal Outcomes in Spain. | 2020 | Martinez-P | Spain | 12/03/2020 | 06/04/2020 | Cohort | 82 | 82 | 78 | 78 | All pregnant women | PCR-confirmed COVID-19 |
| 42 | Vaginal delivery in SARS-CoV-2-infected pregnant women in Northern Italy: a retrospective analysis. | 2020 | Ferrazzi, E | Italy | 01/03/2020 | 20/03/2020 | Cohort | 42 | 42 | 42 | 42 | All pregnant women | PCR-confirmed COVID-19 |
| 43 | Characteristics and Outcomes of 241 Births to Women With Severe Acute Respiratory Syndrome Coronavirus 2 (SARS-CoV-2) Infection at Five New York City Medical Centers. | 2020 | Khoury, Ra | USA | 12/03/2020 | 12/04/2020 | Cohort | 241 | 241 | 245 | 245 | All pregnant women | PCR-confirmed COVID-19 |
| 44 | Evaluating Clinical Course and Risk Factors of Infection and Demographic Characteristics of Pregnant Women with COVID-19 in Hamadan Province, West of Iran. | 2020 | Sattari, Ma | Iran | 06/01/2020 | 21/06/2020 | Cohort | 50 | 50 | 25 | 25 | All pregnant women | COVID-19 diagnosed on clinical criteria |
| 45 | Birth and Infant Outcomes Following Laboratory-Confirmed SARS-CoV-2 Infection in Pregnancy - SET-NET, 16 Jurisdictions, March 29-October 14, 2020. | 2020 | Woodwort | USA | 29/03/2020 | 14/10/2020 | Cohort | 5252 | 5252 | 4495 | 4495 | All pregnant women | Not explicitly stated |

| | | | | | | | | | | | | | |
|---|---|---|---|---|---|---|---|---|---|---|---|---|---|
| 46 | Pregnant women with COVID-19 and risk of adverse birth outcomes and maternal-fetal vertical transmission: a population-based cohort study in Wuhan, China. | 2020 | Yang, Rong | China | 13/01/2020 | 18/03/2020 | Cohort | 65 | 65 | 58 | 58 | All pregnant women | PCR or clinical criteria based diagnosis of COVID-19 |
| 47 | Maternal and neonatal outcomes in COVID-19 infected pregnancies: a prospective cohort study. | 2020 | Pirjani, Rei | Iran | 01/03/2020 | 01/09/2020 | Cohort | 43 | 43 | 43 | 43 | All pregnant women | PCR-confirmed COVID-19 |
| 48 | Neonatal outcome in 29 pregnant women with COVID-19: A retrospective study in Wuhan, China. | 2020 | Wu, Yan-Ti | China | 30/01/2020 | 10/03/2020 | Cohort | 29 | 29 | 29 | 29 | All pregnant women | PCR or radiological diagnosis of COVID-19 |
| 49 | Pregnancy and postpartum outcomes in a universally tested population for SARS-CoV-2 in New York City: a prospective cohort study. | 2020 | Prabhu, M | USA | 24/03/2020 | 20/04/2020 | Cohort | 70 | 70 | 69 | 69 | All pregnant women | PCR-confirmed COVID-19 |
| 50 | SARS-CoV-2 infection among hospitalized pregnant women: reasons for admission and pregnancy characteristics - eight U.S. health care centers, March 1-May 30, 2020. | 2020 | Panagiotak | USA | 01/03/2020 | 30/05/2020 | Cohort | 105 | 105 | 93 | 93 | All hospitalised pregnant women | PCR-confirmed COVID-19 |
| 51 | Characteristics and maternal and birth outcomes of hospitalized pregnant women with laboratory-confirmed COVID-19 - COVID-NET, 13 states, March 1-August 22, 2020. | 2020 | Delahoy, M | USA | 01/03/2020 | 22/08/2020 | Cohort | 458 | 458 | 448 | 448 | All pregnant women | PCR-confirmed COVID-20 |

| | | | | | | | | | | | | | |
|---|---|---|---|---|---|---|---|---|---|---|---|---|---|
| 52 | Safety of vaginal delivery in women infected with COVID-19. | 2021 | Lopian, M. | Israel | 23/03/2020 | 08/05/2020 | Cohort | 21 | 21 | 21 | 21 | All pregnant women | PCR-confirmed COVID-19 |
| 53 | Pregnancy with COVID-19 infection and fetomaternal outcomes. | 2021 | Sharma Ni | India | 21/04/2020 | 07/09/2020 | Cohort | 125 | 125 | 97 | 97 | All pregnant women | PCR-confirmed COVID-19 |
| 54 | Severe acute respiratory syndrome coronavirus 2 (SARS-CoV-2) universal screening in gravids during labor and delivery. | 2021 | Saviron-Co | Spain | 31/03/2020 | 31/08/2020 | Cohort | 6 | 6 | 6 | 6 | All pregnant women | PCR-confirmed COVID-19 |
| 55 | Clinical findings and disease severity in hospitalized pregnant women with coronavirus disease 2019 (COVID-19). | 2020 | Savasi, V. N | Italy | 23/02/2020 | 28/03/2020 | Cohort | 77 | 77 | 57 | 57 | All pregnant women | PCR-confirmed COVID-19 |
| 56 | Neonatal management and outcomes during the COVID-19 pandemic: an observation cohort study. | 2020 | Salvatore, | USA | 22/03/2020 | 17/05/2020 | Cohort | 116 | 116 | 120 | 120 | All pregnant women | PCR-confirmed COVID-19 |
| 57 | Maternal COVID-19 infection, clinical characteristics, pregnancy, and neonatal outcome: a prospective cohort study. | 2020 | Antoun, L. | UK | 01/02/2020 | 01/04/2020 | Cohort | 23 | 23 | 20 | 20 | All pregnant women | PCR-confirmed COVID-19 |
| 58 | Outcomes of maternal-newborn dyads after maternal SARS-CoV-2. | 2020 | Verma, S. a | USA | 01/03/2020 | 10/05/2020 | Cohort | 149 | 149 | 149 | 149 | All pregnant women | PCR-confirmed COVID-19 |
| 59 | Effects of severe acute respiratory syndrome coronavirus 2 infection on pregnant women and their infants: a retrospective study in Wuhan, China. | 2020 | Yang Hui a | China | 20/01/2020 | 19/03/2020 | Cohort | 23 | 23 | 23 | 23 | All pregnant women | PCR-confirmed COVID-19 |
| 60 | Coronavirus disease 2019 in pregnancy was associated with maternal morbidity and preterm birth. | 2020 | Sentilhes, I | France | 01/03/2020 | 03/04/2020 | Cohort | 54 | 54 | 21 | 21 | All pregnant women | PCR or clinical criteria based diagnosis of COVID-19 |

| | | | | | | | | | | | | | |
|---|---|---|---|---|---|---|---|---|---|---|---|---|---|
| 61 | Clinical characteristics of 46 pregnant women with a severe acute respiratory syndrome coronavirus 2 infection in Washington state. | 2020 | Lokken, E. | USA | 21/01/2020 | 17/04/2020 | Cohort | 46 | 46 | 8 | 8 | All pregnant women | PCR-confirmed COVID-19 |
| 62 | SARS-CoV-2/COVID-19 infection in pregnancy and its outcome in a rural tertiary care centre of West Bengal. | 2020 | Saha, M. M | India | Not stated | Not stated | Cohort | 3 | 3 | 3 | 3 | All women attending hospital | PCR-confirmed COVID-19 |
| 63 | The impact of COVID-19 infection on labor and delivery, newborn nursery, and neonatal intensive care unit: prospective observational data from a single hospital system. | 2020 | Griffin, I. a | USA | 21/04/2020 | 05/05/2020 | Cohort | 27 | 27 | 27 | 27 | All pregnant women | PCR or clinical criteria based diagnosis of COVID-19 |
| 64 | Clinical course of coronavirus disease-2019 in pregnancy. | 2020 | Pereira, Au | Spain | 14/03/2020 | 14/04/2020 | Cohort | 60 | 60 | 23 | 23 | All pregnant women | PCR-confirmed COVID-19 |
| 65 | Pregnancy Outcomes in COVID-19: A Prospective Cohort Study in Singapore. | 2020 | Mattar, Cit | Singapore | 15/03/2020 | 22/08/2020 | Cohort | 16 | 16 | 5 | 5 | All pregnant women | PCR-confirmed COVID-19 |
| 66 | A pandemic center's experience of managing pregnant women with COVID-19 infection in Turkey: A prospective cohort study. | 2020 | Sahin, Dile | Turkey | 11/03/2020 | 11/06/2020 | Cohort | 29 | 29 | 10 | 10 | All pregnant women | PCR-confirmed COVID-19 |
| 67 | Vertical Transmission of COVID-19 to the Neonate. | 2020 | Moreno, S | USA | 20/03/2020 | 30/04/2020 | Cohort | 19 | 19 | 21 | 21 | All pregnant women | PCR-confirmed COVID-19 |
| 68 | Maternal and Neonatal Outcomes of Pregnant Women With Coronavirus Disease 2019 (COVID-19) Pneumonia: A Case-Control Study. | 2020 | Li, Na and | China | 24/01/2020 | 29/02/2020 | Cohort | 16 | 16 | 17 | 17 | All pregnant women | PCR-confirmed COVID-19 |

| | | | | | | | | | | | | | |
|---|---|---|---|---|---|---|---|---|---|---|---|---|---|
| 69 | Maternal and perinatal characteristics and outcomes of pregnancies complicated with COVID-19 in Kuwait. | 2020 | Ayed, Ama | Kuwait | 15/03/2020 | 31/05/2020 | Cohort | 185 | 185 | 167 | 167 | All pregnant women | PCR-confirmed COVID-19 |
| 70 | Pregnancy Outcomes Among Women With and Without Severe Acute Respiratory Syndrome Coronavirus 2 Infection. | 2020 | Adhikari, E | USA | 18/03/2020 | 22/08/2020 | Cohort | 252 | 252 | 251 | 251 | All pregnant women | PCR-confirmed COVID-19 |
| 71 | SARS-CoV-2 screening of asymptomatic women admitted for delivery must be performed with a combination of microbiological techniques: an observational study. | 2020 | Vinuela, M | Spain | 06/05/2020 | 21/05/2020 | Cohort | 9 | 9 | 9 | 9 | All pregnant women | PCR-confirmed COVID-19 |
| 72 | Disease severity and perinatal outcomes of pregnant patients with coronavirus disease 2019 (COVID-19). | 2021 | Metz, T. D. | USA | 01/03/2020 | 31/07/2020 | Cohort | 1291 | 1291 | 1196 | 1196 | All pregnant women | PCR-confirmed COVID-19 |
| 73 | Assessing disease outcome in COVID-19 pregnancies in a tertiary referral center in South India: a single-center retrospective cohort study. | 2020 | Nambiar, S | India | 01/04/2020 | 01/09/2020 | Cohort | 350 | 350 | 253 | 254 | All pregnant women | confirmed COVID-19 infection, unclear of method of confirmation |
| 74 | Screening of severe acute respiratory syndrome coronavirus-2 infection during labor and delivery using polymerase chain reaction and immunoglobulin testing. | 2021 | Saviron-Co | Spain | 31/03/2020 | 30/09/2020 | Cohort | 22 | 22 | 22 | 22 | All pregnant women | PCR-confirmed COVID-19 |

| | | | | | | | | | | | | | |
|---|---|---|---|---|---|---|---|---|---|---|---|---|---|
| 75 | Management of gestational diabetes in women with a concurrent severe acute respiratory syndrome coronavirus 2 infection, experience of a single center in northern Italy. | 2020 | D'Ambrosi, | Italy | 01/03/2020 | 30/04/2020 | Cohort | 6 | 6 | 6 | 6 | All pregnant women | PCR-confirmed COVID-19 |
| 76 | Epidemiology, management and risk of SARS-CoV-2 transmission in a cohort of newborns born to mothers diagnosed with COVID-19 infection. | 2021 | Solis-Garci | Spain | 01/03/2020 | 17/08/2020 | Cohort | 73 | 73 | 75 | 75 | All pregnant women | PCR-confirmed COVID-19 |
| 77 | Outcomes of neonates born to mothers with severe acute respiratory syndrome coronavirus 2 infection at a large medical center in New York city. | 2021 | Dumitriu, D | USA | 13/03/2020 | 24/04/2020 | Cohort | 100 | 100 | 101 | 101 | All pregnant women | PCR or clinical criteria based diagnosis of COVID-19 |
| 78 | Neonates born to mothers with COVID-19: data from the Spanish society of neonatology registry. | 2021 | Sanchez-Lu | Spain | 08/03/2020 | 26/05/2020 | Cohort | 493 | 493 | 503 | 503 | All pregnant women | PCR or serology-confirmed COVID-19 |
| 79 | Comparison of hematological parameters and perinatal outcomes between COVID-19 pregnancies and healthy pregnancy cohort. | 2021 | Koc, E. M. | Turkey | 20/03/2020 | 25/07/2020 | Case-control | 39 | 39 | 39 | 39 | All pregnant women admitted | PCR-confirmed COVID-19 |
| 80 | SARS-CoV-2 in pregnancy: maternal and perinatal outcome data of 34 pregnant women hospitalised between may and October 2020. | 2021 | Hall, M. an | Austria | 11/05/2020 | 14/10/2020 | Cohort | 35 | 35 | 28 | 28 | All pregnant women | PCR-confirmed COVID-19 |

| | | | | | | | | | | | | | |
|---|---|---|---|---|---|---|---|---|---|---|---|---|---|
| 81 | The Relationship between Status at Presentation and Outcomes among Pregnant Women with COVID-19. | 2020 | London, Vi | USA | 15/03/2020 | 10/04/2020 | Cohort | 58 | 58 | 55 | 55 | All pregnant women | PCR-confirmed COVID-19 |
| 82 | Analysis of vaginal delivery outcomes among pregnant women in Wuhan, China during the COVID-19 pandemic. | 2020 | Liao, Jing a | China | 20/01/2020 | 02/03/2020 | Case-control | 10 | 10 | 10 | 10 | All pregnant women delivering during study period | COVID-19 diagnosed on clinical criteria |
| 83 | A multicenter study on epidemiological and clinical characteristics of 125 newborns born to women infected with COVID-19 by Turkish Neonatal Society. | 2021 | Oncel, Mel | Turkey | 15/03/2020 | 15/06/2020 | Cohort | 125 | 125 | 120 | 120 | All pregnant women | PCR-confirmed COVID-19 |
| 84 | Assessment of Maternal and Neonatal SARS-CoV-2 Viral Load, Transplacental Antibody Transfer, and Placental Pathology in Pregnancies During the COVID-19 Pandemic. | 2020 | Edlow, And | USA | 02/04/2020 | 13/06/2020 | Cohort | 64 | 64 | 64 | 64 | All pregnant women | PCR-confirmed COVID-19 |
| 85 | Maternal and perinatal outcomes of pregnant women with SARS-CoV-2 infection. | 2021 | WAPM (W | Multiple | 01/02/2020 | 30/04/2020 | Cohort | 388 | 388 | 266 | 266 | All pregnant women | PCR-confirmed COVID-19 |
| 86 | Impact of the Coronavirus Infection in Pregnancy: A Preliminary Study of 141 Patients. | 2020 | Nayak, Aru | India | 01/04/2020 | 15/05/2020 | Cohort | 141 | 141 | 131 | 131 | All pregnant women | PCR-confirmed COVID-19 |
| 87 | [Perinatal COVID-19 in Latin America]. | 2020 | Sola, Augu | Latin Americ | 06/03/2020 | 30/05/2020 | Cohort | 86 | 86 | 86 | 86 | All pregnant women | PCR-confirmed COVID-19 |
| 88 | Clinical characteristics, maternal and neonatal outcomes of pregnant women with SARS-CoV-2 infection in Turkey. | 2021 | Ozsurmeli, | Turkey | 11/03/2020 | 01/07/2020 | Cohort | 24 | 24 | 10 | 10 | All pregnant women | PCR-confirmed COVID-19 |

| | | | | | | | | | | | | | |
|---|---|---|---|---|---|---|---|---|---|---|---|---|---|
| 89 | Maternal, fetal and neonatal outcomes of large series of SARS-CoV-2 positive pregnancies in peripartum period: A single-center prospective comparative study. | 2021 | Hcini, Naje | French Guian | 16/06/2020 | 16/08/2020 | Cohort | 137 | 137 | 127 | 127 | All pregnant women | PCR-confirmed COVID-19 |
| 90 | [Perinatal outcomes and serological results in neonates of pregnant women seropositive to SARS-CoV-2: A cross-sectional descriptive study]. | 2020 | Davila-Alia | Peru | 15/04/2020 | 10/05/2020 | Cohort | 114 | 114 | 114 | 114 | All pregnant women admitted | Serological diagnosis of COVID-19 |
| 91 | Clinical Stratification of Pregnant COVID-19 Patients based on Severity: A Single Academic Center Experience. | 2021 | Berry, Mar | USA | 01/03/2020 | 01/07/2020 | Cohort | 91 | 91 | 60 | 60 | All pregnant women admitted | PCR-confirmed COVID-19 |
| 92 | Pregnancy and neonatal outcomes of COVID-19: coreporting of common outcomes from PAN-COVID and AAP-SONPM registries. | 2021 | Mullins, E | Worldwide | 01/01/2020 | 25/07/2020 | Cohort | 3050 | 3050 | 3050 | 3050 | All pregnant women | PCR-confirmed COVID-19 |
| 93 | Updated experience of a tertiary pandemic center on 533 pregnant women with COVID-19 infection: A prospective cohort study from Turkey. | 2021 | Sahin, Dile | Turkey | 11/03/2020 | 10/09/2020 | Cohort | 533 | 533 | 131 | 131 | All pregnant women | PCR-confirmed COVID-19 |
| 94 | Maternal and Neonatal Outcomes of COVID-19 in Pregnancy: A Single-Centre Observational Study. | 2021 | Singh, Vini | India | 15/05/2020 | 15/11/2020 | Cohort | 132 | 132 | 125 | 125 | All pregnant women | PCR-confirmed COVID-19 |

| | | | | | | | | | | | | | |
|---|---|---|---|---|---|---|---|---|---|---|---|---|---|
| 95 | Characteristics and outcomes of neonatal SARS-CoV-2 infection in the UK: a prospective national cohort study using active surveillance. | 2021 | Gale, Chris | UK | 01/03/2020 | 30/04/2020 | Cohort | 17 | 17 | 66 | 66 | All neonates with confirmed SARS-CoV-2 | PCR-confirmed COVID-19 |
| 96 | COVID-19 in a cohort of pregnant women and their descendants, the MOACC-19 study. | 2021 | Llorca, Javi | Spain | 23/05/2020 | 22/10/2020 | Cohort | 14 | 14 | 14 | 14 | All pregnant women | PCR-confirmed COVID-19 |
| 97 | Coronavirus disease 2019 in pregnancy | 2020 | Qiancheng | China | 15/01/2020 | 15/03/2020 | Cohort | 82 | 82 | 23 | 23 | All pregnant women | PCR-confirmed COVID-19 |
| 98 | Clinical analysis of ten pregnant women with COVID-19 in Wuhan, China: A retrospective study | 2020 | Cao, Dong | China | 23/01/2020 | 23/02/3030 | Cohort | 10 | 10 | 10 | 10 | All pregnant women | PCR-confirmed COVID-19 |
| 99 | Maternal, Perinatal and Neonatal Outcomes with COVID-19: A Multicenter Study of 242 Pregnancies and Their 248 Infant Newborns during Their First Month of Life | 2020 | Reyne Ver | Spain | 13/03/2020 | 31/05/2020 | Cohort | 242 | 242 | 248 | 248 | All pregnant women | PCR or serology-confirmed COVID-19 |
| 100 | Clinical course of novel COVID-19 infection in pregnant women | 2020 | Shmakov, I | Russia | Not stated | Not stated | Cohort | 66 | 66 | 42 | 42 | All pregnant women admitted | PCR-confirmed COVID-19 |
| 101 | Vaginal delivery in SARS-CoV-2-infected pregnant women in Israel: a multicenter prospective analysis | 2020 | Rottenstre | Israel | 15/03/2020 | 04/07/2020 | Cohort | 52 | 52 | 52 | 52 | All pregnant women | PCR-confirmed COVID-19 |
| 102 | Clinical Analysis of Neonates Born to Mothers with or without COVID-19: A Retrospective Analysis of 48 Cases from Two Neonatal Intensive Care Units in Hubei Province | 2020 | Liu, Wei ar | China | 17/01/2020 | 04/03/2020 | Case-control | 31 | 15 | 31 | 15 | All pregnant women | PCR or radiological diagnosis of COVID-19 |

| | | | | | | | | | | | | | |
|---|---|---|---|---|---|---|---|---|---|---|---|---|---|
| 103 | Characteristics of Newborns Born to SARS-CoV-2-Positive Mothers: A Retrospective Cohort Study | 2020 | Kupfermar | USA | 01/03/2020 | 01/05/2020 | Case-control | 79 | 15 | 79 | 15 | All pregnant women | PCR-confirmed COVID-19 |
| 104 | Poor maternal-neonatal outcomes in pregnant patients with confirmed SARS-Cov-2 infection: analysis of 145 cases | 2021 | Di Guardo, | Italy | 01/03/2020 | 01/07/2020 | Cohort | 145 | 145 | 145 | 145 | All pregnant women | PCR-confirmed COVID-19 |
| 105 | Coronavirus and birth in Italy: results of a national population-based cohort study | 2020 | Maraschin | Italy | 25/02/2020 | 22/04/2020 | Cohort | 146 | 146 | 147 | 147 | All pregnant women | PCR, serological or radiological diagnosis of COVID-19 |
| 106 | Infant Outcomes Following Maternal Infection with SARS-CoV-2: First Report from the PRIORITY Study | 2020 | Flaherman | USA | 22/03/2020 | 22/06/2020 | Case-control | 263 | 179 | 263 | 179 | All pregnant women | PCR-confirmed COVID-19 |
| 107 | Incidence and clinical profiles of COVID-19 pneumonia in pregnant women: A single-centre cohort study from Spain | 2020 | San-Juan, I | Spain | 05/03/2020 | 05/04/2020 | Cohort | 52 | 52 | 6 | 6 | All pregnant women | PCR-confirmed COVID-19 |
| 108 | Vaginal delivery in SARS-CoV-2-infected pregnant women in Northern Italy: a retrospective analysis | 2020 | Iurlaro, E. a | Italy | 01/03/2020 | 20/03/2020 | Cohort | 42 | 42 | 42 | 42 | All pregnant women | PCR-confirmed COVID-19 |
| 109 | Obstetric Outcomes of SARS-CoV-2 Infection in Asymptomatic Pregnant Women | 2021 | Cruz-Lemir | Spain | 23/03/2020 | 31/03/2020 | Case-control | 604 | 174 | 604 | 174 | All pregnant women | PCR-confirmed COVID-19 |
| 110 | SARS-CoV-2 infection in pregnancy and newborn in a Spanish multicentric cohort (GESNEO-COVID) | 2020 | Carrasco, I | Spain | 15/03/2020 | 31/07/2020 | Cohort | 105 | 105 | 107 | 107 | All pregnant women | PCR or serology-confirmed COVID-19 |

| | | | | | | | | | | | | | |
|---|---|---|---|---|---|---|---|---|---|---|---|---|---|
| 111 | Clinical course of severe and critical coronavirus disease 2019 in hospitalized pregnancies: a United States cohort study | 2020 | Pierce-Will | USA | 05/03/2020 | 20/04/2020 | Cohort | 64 | 64 | 33 | 33 | All pregnant women | PCR-confirmed COVID-19 |
| 112 | Rates of seroprevalance of COVID-19 among pregnant patients in New York City | 2021 | Baptiste, C | USA | 24/06/2020 | 16/07/2020 | Cohort | 47 | 19 | 47 | 19 | All pregnant women | PCR or serology-confirmed COVID-19 |
| 113 | Maternal and neonatal outcomes of pregnant patients with coronavirus disease 2019 (COVID-19): A multistate cohort | 2021 | Metz, Torr | USA | 01/03/2020 | 31/07/2020 | Cohort | 1219 | 1219 | 1196 | 1196 | All pregnant women | PCR or serology-confirmed COVID-19 |
| 114 | Obstetrical and neonatal outcomes among pregnancies with SARS-CoV-2 | 2021 | Trahan, M | Canada | 22/03/2020 | 31/07/2020 | Case-control | 206 | 43 | 209 | 45 | All pregnant women | PCR-confirmed COVID-19 |
| 115 | Comparison of clinical outcomes in pregnant women with and without COVID-19 based on disease severity | 2021 | Gold, Stace | USA | 01/03/2020 | 01/07/2020 | Case-control | 486 | 91 | 486 | 91 | All pregnant women delivering during study period | Not explicitly stated |
| 116 | Neonatal outcomes of COVID-19 positive mothers | 2021 | Zarudskaya | USA | 01/03/2020 | 01/08/2020 | Case-control | 28 | 10 | 28 | 10 | All pregnant women | PCR-confirmed COVID-19 |
| 117 | Perinatal outcomes of asymptomatic versus symptomatic COVID positive pregnant women | 2021 | Andrikopo | USA | 12/03/2020 | 12/08/2020 | Cohort | 157 | 157 | 156 | 156 | All pregnant women | PCR-confirmed COVID-19 |
| 118 | Neonatal outcomes in pregnant women with diagnosis of COVID-19 | 2021 | Izewski, Jo | USA | 01/03/2020 | 31/03/2020 | Case-control | 515 | 460 | 55 | 55 | All pregnant women | PCR-confirmed COVID-19 |
| 119 | Infant outcomes and maternal COVID-19 status at delivery | 2021 | Cohen, Lou | USA | 15/03/2020 | 15/06/2020 | Case-control | 184 | 60 | 186 | 62 | All pregnant women | PCR-confirmed COVID-19 |
| 120 | Initial review of pregnancy and neonatal outcomes of pregnant women with COVID-19 infection | 2021 | Ogamba, If | USA | 17/03/2020 | 04/06/2020 | Cohort | 40 | 40 | 25 | 25 | All pregnant women | PCR or serology-confirmed COVID-19 |

| | | | | | | | | | | | | | |
|---|---|---|---|---|---|---|---|---|---|---|---|---|---|
| 121 | Impact of SARS-CoV-2 Infection on Pregnancy Outcomes: A Population-Based Study | 2021 | Larroya, M | Spain | 15/03/2020 | 31/05/2020 | Case-control | 2225 | 317 | 1338 | 178 | All pregnant women | PCR or serology-confirmed COVID-19 |
| 122 | Management and short-term outcomes of infants born to mothers with active perinatal covid-19 infection | 2021 | Lien, J. and | USA | 01/04/2020 | 01/10/2020 | Cohort | 43 | 43 | 43 | 43 | All pregnant women | PCR-confirmed COVID-19 |
| 123 | Maternal and perinatal outcomes of pandemic Covid-19 in pregnancy in Basrah | 2021 | Sharief, M | Iraq | 15/03/2020 | 01/11/2020 | Cohort | 135 | 135 | 110 | 110 | All pregnant women | PCR-confirmed COVID-19 |
| 124 | Maternal, neonatal and placental characteristics of SARS-CoV-2 positive mothers | 2021 | Zhang, Pei | USA | 01/03/2020 | 01/08/2020 | Case-control | 219 | 142 | 219 | 142 | All pregnant women | PCR-confirmed COVID-19 |
| 125 | Disease severity, pregnancy outcomes, and maternal deaths among pregnant patients with severe acute respiratory syndrome coronavirus 2 infection in Washington State | 2021 | Lokken, Eri | USA | 01/03/2020 | 30/06/2020 | Cohort | 240 | 240 | 156 | 156 | All pregnant women | PCR-confirmed COVID-19 |
| 126 | Maternal and perinatal outcomes in high vs low risk-pregnancies affected by SARS-COV-2 infection (Phase-2): The WAPM (World Association of Perinatal Medicine) working group on COVID-19 | 2021 | D'Antonio, | 25 countries | 04/04/2020 | 28/10/2020 | Cohort | 887 | 887 | 874 | 874 | All pregnant women | PCR-confirmed COVID-19 |

| | | | | | | | | | | | | | |
|---|---|---|---|---|---|---|---|---|---|---|---|---|---|
| 127 | Epidemiology of coronavirus disease 2019 in pregnancy: risk factors and associations with adverse maternal and neonatal outcomes | 2021 | Brandt, Jus | USA | 11/03/2020 | 11/06/2020 | Case-control | 183 | 122 | 184 | 123 | All pregnant women | PCR-confirmed COVID-19 |
| 128 | Covid 19 infection in pregnant women and newborn infants at a single U.S. center: What disparities, testing and isolation practices can teach US | 2020 | Camelo, In | USA | 31/03/2020 | 17/06/2020 | Cohort | 36 | 36 | 32 | 32 | All pregnant women | PCR-confirmed COVID-19 |
| 129 | The association between SARS-CoV-2 infection and preterm delivery: a prospective study with a multivariable analysis | 2021 | Martinez-P | Spain | 23/03/2020 | 31/05/2020 | Case-control | 1009 | 246 | 1009 | 246 | All pregnant women | PCR-confirmed COVID-19 |
| 130 | Pregnancy Outcomes among Women with and without Severe Acute Respiratory Syndrome Coronavirus 2 Infection | 2020 | McIntire, D | USA | 18/03/2020 | 22/08/2020 | Case-control | 3374 | 252 | 3263 | 248 | All pregnant women | PCR-confirmed COVID-19 |
| 131 | Clinicolaboratory profile of and outcomes in neonates born to covid-19-positive mothers | 2021 | Sehra, Ram | India | 13/04/2020 | 31/07/2020 | Cohort | 120 | 120 | 120 | 120 | All pregnant women | PCR-confirmed COVID-19 |
| 132 | Clinical Profile, Viral Load, Maternal-Fetal Outcomes of Pregnancy With COVID-19: 4-Week Retrospective, Tertiary Care Single-Centre Descriptive Study | 2021 | Bachani, Su | India | 05/05/2020 | 05/06/2020 | Cohort | 57 | 57 | 56 | 56 | All pregnant women | PCR-confirmed COVID-19 |
| 133 | Management and Early Outcomes of Neonates Born to Women with SARS-CoV-2 in 16 US. Hospitals | 2021 | Congdon, J | USA | 01/03/2020 | 01/05/2020 | Cohort | 70 | 70 | 70 | 70 | All pregnant women | PCR-confirmed COVID-19 |

| | | | | | | | | | | | | | |
|---|---|---|---|---|---|---|---|---|---|---|---|---|---|
| 134 | Maternal and neonatal outcomes of pregnant patients with COVID-19: A prospective cohort study | 2021 | Abedzadeh | Iran | 01/03/2020 | 01/11/2020 | Case-control | 150 | 56 | 149 | 55 | All pregnant women | PCR-confirmed COVID-19 |
| 135 | Multicentre Spanish study found no incidences of viral transmission in infants born to mothers with COVID-19 | 2020 | Marin Gab | Spain | 13/03/2020 | 29/03/2020 | Cohort | 42 | 42 | 42 | 42 | All pregnant women | PCR-confirmed COVID-19 |
| 136 | SARS-CoV-2 Infection during Pregnancy in a Rural Midwest All-delivery Cohort and Associated Maternal and Neonatal Outcomes | 2021 | Steffen, Ha | USA | 01/05/2020 | 22/09/2020 | Case-control | 1000 | 61 | 1021 | 62 | All pregnant women | PCR-confirmed COVID-19 |
| 137 | Maternal and perinatal outcomes of pregnant women with SARS-CoV-2 infection | 2021 | WAPM Wo | Multiple | 01/02/2020 | 30/04/2020 | Cohort | 388 | 388 | 251 | 251 | All pregnant women | PCR-confirmed COVID-19 |
| 138 | A single-center observational study on clinical features and outcomes of 21 SARS-CoV-2-infected neonates from India | 2021 | Nanavati, F | India | 15/04/2020 | 31/07/2020 | Cohort | 122 | 122 | 125 | 125 | All pregnant women | PCR-confirmed COVID-19 |
| 139 | Perinatal outcomes in pregnant women with COVID-19 in Siberia and the Russian Far East | 2021 | Artymuk, N | Russia | Not stated | 25/12/2020 | Cohort | 8485 | 8485 | 2383 | 2383 | All pregnant women | PCR-confirmed COVID-20 |
| 140 | Prevalence and Risk Factors of Neonatal Covid-19 Infection: A Single-Centre Observational Study | 2021 | Ajith, S. an | India | 15/04/2020 | 15/10/2020 | Cohort | 350 | 350 | 223 | 223 | All pregnant women | PCR-confirmed COVID-19 |

| | | | | | | | | | | | | | |
|---|---|---|---|---|---|---|---|---|---|---|---|---|---|
| 141 | Impact of Covid-19 in pregnancy on mother's psychological status and infant's neurobehavioral development: a longitudinal cohort study in China. | 2020 | Wang, Yua | China | 01/05/2020 | 31/07/2020 | Cohort | 72 | 72 | 57 | 57 | All pregnant women | PCR-confirmed COVID-19 |
| 142 | Vertical Transmission of Novel Coronavirus (COVID-19) from Mother to Newborn: Experience from a Maternity Unit, The Indus Hospital, Karachi. | 2020 | Khan, Muh | Pakistan | 27/04/2020 | 16/06/2020 | Cohort | 66 | 66 | 67 | 67 | All pregnant women | PCR-confirmed COVID-19 |
| 143 | Coronavirus disease 2019 in pregnant women: a report based on 116 cases. | 2020 | Yan Jie and | China | 20/01/2020 | 24/03/2020 | Cohort | 116 | 116 | 100 | 100 | All pregnant women | PCR or clinical criteria based diagnosis of COVID-19 |
| 144 | Clinical profile of SARS-CoV-2 infected neonates from a tertiary government hospital in Mumbai, India. | 2020 | Pavan Kala | India | 01/04/2020 | 31/05/2020 | Cohort | 185 | 185 | 185 | 185 | All pregnant women | PCR-confirmed COVID-19 |
| 145 | Clinical profile, viral load, management and outcome of neonates born to COVID 19 positive mothers: a tertiary care centre experience from India. | 2020 | Pratima Ar | India | 01/04/2020 | 10/07/2020 | Cohort | 69 | 69 | 65 | 65 | All pregnant women | PCR-confirmed COVID-19 |
| 146 | Clinical Manifestation and Neonatal Outcomes of Pregnant Patients With Coronavirus Disease 2019 Pneumonia in Wuhan, China. | 2020 | Xu, Shuang | China | 15/01/2020 | 15/03/2020 | Cohort | 64 | 64 | 23 | 23 | All pregnant women | PCR-confirmed COVID-19 |
| 147 | Impact of SARS-CoV-2 on multiple gestation pregnancy. | 2021 | Mahajan, N | India | 04/04/2020 | 10/09/2020 | Cohort | 879 | 879 | 633 | 633 | All pregnant women | PCR-confirmed COVID-19 |

| | | | | | | | | | | | | | |
|---|---|---|---|---|---|---|---|---|---|---|---|---|---|
| 148 | The impact of perinatal severe acute respiratory syndrome coronavirus 2 infection during the peripartum period. | 2021 | Janssen, O | USA | 25/03/2020 | 15/05/2020 | Cohort | 180 | 180 | 180 | 180 | All pregnant women | PCR-confirmed COVID-19 |
| 149 | Maternal and perinatal characteristics of pregnant women with COVID-19 in a national hospital in Lima, Peru | 2020 | Huerta Sae | Peru | 24/03/2020 | 07/05/2020 | Cohort | 37 | 37 | 35 | 35 | All pregnant women | PCR or rapid test confirmed COVID-19 |
| 150 | An initiative to evaluate the safety of maternal bonding in patients with SARS-CoV-2 infection | 2020 | Cojocaru, L | USA | 01/03/2020 | 01/06/2020 | Cohort | 86 | 86 | 34 | 34 | All pregnant women | PCR-confirmed COVID-19 |
| 151 | Short-term neonatal outcomes of colocating and breastfeeding infants of mothers who tested positive for sars-cov-2 | 2020 | Krishnan, P | USA | 19/03/2020 | 22/04/2020 | Cohort | 45 | 45 | 45 | 45 | All pregnant women | PCR-confirmed COVID-19 |
| 152 | Impact of Maternal Severe Acute Respiratory Syndrome Coronavirus 2 Detection on Breastfeeding Due to Infant Separation at Birth | 2020 | Popofsky, S | USA | 25/03/2020 | 30/05/2020 | Cohort | 160 | 160 | 160 | 160 | All pregnant women | PCR-confirmed COVID-19 |
| 153 | SARS-CoV-2 in pregnancy: characteristics and outcomes of hospitalized and non-hospitalized women due to COVID-19 | 2020 | Barbero, P | Spain | 03/03/2020 | 31/05/2020 | Cohort | 23 | 23 | 23 | 23 | All pregnant women | PCR-confirmed COVID-19 |
| 154 | Clinical characteristics of COVID-19 in pregnant women: A retrospective descriptive single-center study from a tertiary hospital in Muscat, Oman | 2021 | Santhosh, J | Oman | 24/03/2020 | 31/07/2020 | Cohort | 60 | 60 | 46 | 46 | All pregnant women | PCR-confirmed COVID-19 |

| | | | | | | | | | | | | | |
|---|---|---|---|---|---|---|---|---|---|---|---|---|---|
| 155 | Clinical characteristics and pregnancy outcomes of women diagnosed with SARS-CoV-2 in London's most ethnically diverse borough: A cross-sectional study | 2021 | Milln, Jack | UK | 12/03/2020 | 22/04/2020 | Cohort | 32 | 32 | 30 | 30 | All pregnant women | PCR-confirmed COVID-19 |
| 156 | Evaluation of cochlear functions in infants exposed to SARS-CoV-2 intrauterine | 2021 | Celik, Turg | Turkey | 01/03/2020 | 01/12/2020 | Cohort | 73 | 37 | 73 | 37 | All pregnant women | PCR-confirmed COVID-19 |
| 157 | Ultrasound and Doppler findings in pregnant SARS-CoV-2 positive women | 2021 | Soto-Torre | USA | 01/05/2020 | 31/08/2020 | Cohort | 209 | 106 | 209 | 106 | All pregnant women | PCR or rapid test confirmed COVID-19 |
| 158 | Maternal and perinatal outcomes of pregnant women with SARS-CoV-2 infection at the time of birth in England: national cohort study | 2021 | Gurol-Urga | UK | 29/05/2020 | 31/01/2021 | Case-control | 342080 | 3527 | 338553 | 3527 | All pregnant women delivering singletons | PCR-confirmed COVID-19 |
| 159 | Clinical characteristics of pregnant women with COVID-19 in Wuhan, China | 2020 | Lian Chen | China | 08/12/2019 | 20/03/2020 | Cohort | 118 | 118 | 70 | 70 | All pregnant women | radiological diagnosis of COVID-19 |
| 160 | Clinical features and the maternal and neonatal outcomes of pregnant women with coronavirus 2019 | 2020 | Rui Nie | China | 01/01/2020 | 01/02/2020 | Cohort | 33 | 33 | 28 | 28 | All pregnant women | PCR-confirmed COVID-19 |
| 161 | Coronavirus disease 2019 among pregnant Chinese women: case series data on the safety of vaginal birth and breastfeeding | 2020 | Y Wu | China | 31/01/2020 | 09/03/2020 | Cohort | 13 | 13 | 5 | 5 | All pregnant women admitted | PCR-confirmed COVID-19 |

| | | | | | | | | | | | | | |
|---|---|---|---|---|---|---|---|---|---|---|---|---|---|
| 162 | Severe acute respiratory syndrome coronavirus 2 (SARS-CoV-2) infection during pregnancy in China: a retrospective cohort study | 2020 | Ming-Zhu Y | China | 28/01/2020 | 28/02/2020 | Cohort | 31 | 31 | 17 | 17 | All female patients | PCR-confirmed COVID-19 |
| 163 | Prevalence of SARS-CoV-2 among patients admitted for childbirth in southern Conneticut | 2020 | Katherine ( | USA | 02/04/2020 | 29/04/2020 | Cohort | 770 | 30 | 770 | 30 | All pregnant women admitted for childbirth | PCR-confirmed COVID-19 |
| 164 | Clinical characteristics of pregnant women with Coronavirus disease 2019 in Wuhan, China | 2020 | Biheng Che | China | 15/01/2020 | 23/02/2020 | Cohort | 111 | 111 | 17 | 17 | All pregnant women admitted | PCR-confirmed COVID-19 |
| 165 | COVID-19 during pregnancy: A case series from an universally tested population from the north of Portugal | 2020 | Mariana D | Portugal | 25/03/2020 | 15/04/2020 | Cohort | 12 | 12 | 11 | 11 | All pregnant women admitted | PCR-confirmed COVID-19 |
| 166 | Routine screening for SARS-CoV-2 in unselected pregnant women at delivery | 2020 | Pilar Diaz-( | Chile | 27/04/2020 | 07/06/2020 | Case-control | 583 | 37 | 586 | 37 | All pregnant women admitted | PCR-confirmed COVID-19 |
| 167 | COVID-19 in the second half of pregnancy: prevalence and clinical relevance | 2020 | Marta Rug | Italy | 07/04/2020 | 06/05/2020 | Case-control | 315 | 28 | 315 | 28 | All pregnant women delivering during study period | PCR-confirmed COVID-19 |
| 168 | Racial-ethnic disparities and pregnancy outcomes in SARS-CoV-2 infection in a universally-tested cohort in Houston, Texas | 2020 | Beth Pinele | USA | 22/04/2020 | 22/07/2020 | Case-control | 935 | 77 | 935 | 77 | All pregnant women admitted for childbirth | PCR-confirmed COVID-19 |
| 169 | Fetomaternal outcome in COVID-19 infected pregnant women: a preliminary clinical study | 2020 | Parul Shah | India | 15/04/2020 | 10/06/2020 | Cohort | 125 | 125 | 96 | 96 | All pregnant women | PCR-confirmed COVID-19 |

| | | | | | | | | | | | | |
|---|---|---|---|---|---|---|---|---|---|---|---|---|
| 170 | COVID-19 infection during pregnancy - maternal and perinatal outcomes: a tertiary care centre study | 2020 | Nazia Hass | India | 01/03/2020 | 30/06/2020 | Cohort | 38 | 38 | 37 | 37 | All pregnant women in third trimester | PCR-confirmed COVID-19 |
| 171 | A study of breastfeeding practices, SARS-CoV-2 and its antibodies in the breast milk of mothers confirmed with COVID-19 | 2020 | Sicong Pen | China | 10/02/2020 | 01/04/2020 | Case-control | 64 | 24 | 66 | 25 | All pregnant women | PCR-confirmed COVID-19 |
| 172 | Influence of race and ethnicity on severe acute respiratory syndrome coronavirus 2 (SARS-CoV-2) infectino rates and clinical outcomes in pregnanct | 2020 | Ukachi Em | USA | 13/03/2020 | 23/04/2020 | Cohort | 100 | 100 | 100 | 100 | All pregnant women delivering during study period | PCR-confirmed COVID-19 |
| 173 | Vertical transmission and materno-fetal outcomes in 13 patients with coronavirus disease 2019 | 2020 | S Masmeja | Switzerland | 01/04/2020 | 06/05/2020 | Cohort | 13 | 13 | 13 | 13 | All pregnant women admitted | PCR-confirmed SARS-CoV-2 or serology and positive contact |
| 174 | Retrospective description of pregnant women infected with severe acute respiratory syndrome coronavirus 2, France | 2020 | Alexandre | France | 12/03/2020 | 12/04/2020 | Cohort | 100 | 100 | 36 | 36 | All pregnant women | PCR-confirmed SARS-CoV-2 |
| 175 | Maternal and Fetal Outcomes of Pregnant Women Infected with Coronavirus Based on Tracking the Results of 90-Days Data in Hazrat -E- Rasoul Akram Hospital, Iran University of Medical Sciences | 2021 | Shahla Cha | Iran | 08/03/2020 | 28/12/2020 | Cohort | 14 | 14 | 13 | 13 | All pregnant women hospitalised | PCR-confirmed SARS-CoV-2 and/or typical radiological features |

| | | | | | | | | | | | | | |
|---|---|---|---|---|---|---|---|---|---|---|---|---|---|
| 176 | Clinical Characteristics, Management, and Short-Term Outcome of Neonates Born to Mothers with COVID-19 in a Tertiary Care Hospital in India | 2021 | Sushma M | India | 14/04/2020 | 31/07/2020 | Cohort | 514 | 514 | 524 | 524 | All pregnant women delivering during study period | PCR-confirmed SARS-CoV-2 |
| 177 | Covid-19 and pregnancy: the experience of a tertiary maternity hospital | 2021 | Panagiotis | Greece | 01/03/2020 | 31/12/2020 | Cohort | 40 | 40 | 35 | 35 | All pregnant women admitted | PCR-confirmed SARS-CoV-2 |
| 178 | Neonatal Outcomes in Pregnant Women Infected with COVID-19 in Babol, North of Iran: A Retrospective Study with Short-Term Follow-Up | 2021 | Zahra Akba | Iran | 10/02/2020 | 20/05/2020 | Cohort | 8 | 8 | 8 | 8 | All pregnant women admitted for childbirth | PCR-confirmed SARS-CoV-2 |
| 179 | MATERNAL-PERINATAL OUTCOMES IN PREGNANT WOMEN WITH COVID-19 IN A LEVEL III HOSPITAL IN PERU | 2021 | Carmen Da | Peru | 01/04/2020 | 30/06/2020 | Cohort | 43 | 43 | 43 | 43 | All pregnant women delivering during study period | PCR-confirmed SARS-CoV-2 |
| 180 | Neonatal outcomes of pregnant women with COVID-19 in a developing country setup | 2021 | Manas Kur | India | 01/05/2020 | 20/10/2020 | Cohort | 162 | 162 | 165 | 165 | All pregnant women delivering during study period | PCR-confirmed SARS-CoV-2 |
| 181 | Maternal and Neonatal | 2021 | Moushmi F | India | 01/05/2020 | 31/08/2020 | Case-control | 362 | 181 | 362 | 187 | All pregnant | PCR-confirmed |
| 182 | Pregnancy Outcomes and SARS- | 2021 | Sara Cruz M | Spain | 26/02/2020 | 05/11/2020 | Case-control | 2954 | 1347 | 2954 | 1347 | All pregnant | PCR-confirmed |
| 183 | Placental pathology in COVID- | 2021 | Chiara Tas | Italy | 01/03/2020 | 31/08/2020 | Case-control | 128 | 64 | 128 | 64 | All pregnant | PCR-confirmed |
| 184 | Obstetric, maternal, and | 2021 | Seyed-Abd | Iran | 01/03/2020 | 30/11/2020 | Case-control | 110 | 55 | 110 | 55 | All pregnant | PCR-confirmed |
| 185 | Vertical transmission of | 2021 | Mariane de | Brazil | 12/04/2020 | 30/09/2020 | Cohort | 109 | 109 | 109 | 109 | All pregnant | PCR-confirmed |
| 186 | Consequences of Early | 2021 | Maria Giuli | Italy | 01/04/2020 | 18/03/2021 | Cohort | 37 | 37 | 37 | 37 | All pregnant | PCR-confirmed |
| 187 | Perinatal outcome and possible | 2021 | Ritu Sharm | India | 01/04/2020 | 31/08/2020 | Cohort | 41 | 41 | 44 | 44 | All pregnant | PCR-confirmed |
| 188 | The Relationship between | 2020 | Viktoriya L | USA | 15/03/2020 | 10/04/2020 | Cohort | 55 | 55 | 55 | 55 | All pregnant | PCR-confirmed |
| 189 | Association of SARS-CoV-2 Test | 2020 | Mia Ahlber | Sweden | 25/03/2020 | 24/07/2020 | Case-control | 759 | 156 | 759 | 156 | All pregnant | PCR-confirmed |
| 190 | Infants Born to Mothers with | 2020 | C Murphy | Ireland | 01/03/2020 | 01/07/2020 | Cohort | 26 | 26 | 26 | 26 | All pregnant | PCR-confirmed |
| 191 | Characteristics and outcomes | 2021 | Maryam Va | Iran | 01/03/2020 | 31/10/2020 | Cohort | 110 | 110 | 51 | 51 | All pregnant | PCR-confirmed |
| 192 | Comparing Infection Profiles of | 2021 | Yolanda Cu | Spain | 01/03/2020 | 03/11/2020 | Cohort | 1295 | 1295 | 1295 | 1295 | All pregnant | PCR-confirmed |
| 193 | Coronavirus disease 2019 in | 2021 | Neha Agar | India | 15/04/2020 | 30/06/2020 | Cohort | 65 | 65 | 48 | 48 | All pregnant | PCR-confirmed |
| 194 | COVID-19 infection in | 2021 | Tuba Dama | Turkey | 01/04/2020 | 31/08/2020 | Cohort | 75 | 75 | 34 | 34 | All pregnant | PCR-confirmed |

| | | | | | | | | | | | | | |
|---|---|---|---|---|---|---|---|---|---|---|---|---|---|
| 195 | Maternal and perinatal | 2021 | Ipek Gurol- | England | 29/05/2020 | 31/01/2021 | Case-control | 342080 | 3527 | 302011 | 2555 | All pregnant | PCR-confirmed |
| 196 | Neonatal outcome following | 2021 | Nadine Ma | Germany, Au | 03/04/2020 | 27/11/2020 | Cohort | 435 | 435 | 435 | 435 | All pregnant | PCR-confirmed |
| 197 | Ocular Assessments of a Series | 2021 | Olívia Pere | Brazil | 01/04/2020 | 30/11/2020 | Cohort | 165 | 165 | 165 | 165 | All pregnant | PCR-confirmed |
| 198 | Rooming-in, Breastfeeding and | 2021 | Isabel Brito | Portugal | 01/04/2020 | 31/12/2020 | Cohort | 77 | 77 | 77 | 77 | All pregnant | PCR-confirmed |
| 199 | Perinatal Prognosis in third | 2020 | Denise Veg | Chile | 18/04/2020 | 26/06/2020 | Cohort | 9 | 9 | 9 | 9 | All pregnant | PCR-confirmed |
| 200 | Premature delivery in COVID- | 2020 | Sebastian V | Chile | 17/04/2020 | 30/06/2020 | Case-control | 597 | 59 | 597 | 59 | All pregnant | PCR-confirmed |
| 201 | COVID-19 and pregnancy in | 2020 | Hernandez | Chile | 07/04/2020 | 06/07/2020 | Cohort | 661 | 661 | 316 | 316 | All pregnant | PCR-confirmed |
| 202 | Gestation in times of COVID-19 | 2020 | Vera Loyol | Peru | 01/04/2020 | 01/07/2020 | Cohort | 2214 | 345 | 2214 | 349 | All pregnant | PCR-confirmed |
| 203 | SARS-CoV-2 in the second half | 2020 | Munoz Tay | Peru | 01/03/2020 | 01/07/2020 | Cohort | 247 | 247 | 250 | 250 | All pregnant | PCR-confirmed |
| 204 | SARS-CoV-2 pandemic and | 2020 | Morales | Chile | 01/04/2020 | 01/06/2020 | Cohort | 409 | 78 | 78 | 78 | All pregnant | PCR-confirmed |
